# Artificial Intelligence-Enhanced Electrocardiogram Models for Detection of Left Ventricular Dysfunction: A Comparison Study

**DOI:** 10.1101/2025.07.08.25331129

**Authors:** Philip M. Croon, Machteld J. Boonstra, Cornelis P. Allaart, Bauke K.O. Arends, Lovedeep S. Dhingra, Yu-Chang Huang, Thomas Mast, Rohan Khera, Chang-Fu Kuo, Joon-Myoung Kwon, Hak-Seung Lee, Min Sung Lee, Rutger R. van de Leur, Zhi-Yong Liu, Evangelos K. Oikonomou, Jasper L. Selder, Michiel M. Winter, Folkert W. Asselbergs

## Abstract

**Background:** Several artificial intelligence-enhanced electrocardiogram (AI-ECG) models have shown promise in detecting left ventricular systolic dysfunction (LVSD), but their head-to-head agreement and performance have not been independently compared within the same cohort.

**Objectives:** To compare the performance of published AI-ECG models for LVSD detection in a standardized external cohort and evaluate the field’s transparency and reproducibility.

**Methods:** We systematically reviewed AI-ECG models predicting LVSD and assessed the risk of bias. Authors were invited to share models for external validation in a well-phenotyped registry of patients undergoing routine clinical cardiac magnetic resonance imaging (CMR) with cardiologist-adjudicated reports and paired ECGs. Model performance was evaluated in all consecutive patients and a lower-complexity subgroup with 15% LVSD prevalence.

**Results:** We identified 35 studies describing 51 models, reporting high (AUROC >0.80) or excellent (AUROC >0.90) performance. The risk of bias is high and primarily attributed to the limited description of development and validation cohort characteristics, as well as the lack of independent external validation. Four groups (from Korea, the United States, Taiwan, and the Netherlands) shared models for independent testing. AUROCs ranged from 0.83 to 0.93 in all patients (n = 1,203; mean age 59 ± 15 years; 450 [35%] female) and from 0.87 to 0.96 in the lower complexity subset. Performance remained consistent across subgroups, with slight decreases in ECGs showing wide QRS complexes or atrial fibrillation.

**Conclusions:** In this first-in-kind independent validation and head-to-head comparison study, AI-ECG for LVSD detection demonstrated strong performance despite training on disparate populations. However, the limited availability of models hinders independent validation.

## Introduction

Recent advances in artificial intelligence (AI) have enabled the detection of subtle abnormalities in electrocardiograms (ECGs) that often elude expert interpretation.^1,2^ This expands the ECG’s utility to detect conditions that typically require advanced imaging.^3–7^ Given its low cost and widespread availability, AI-enhanced ECG (AI-ECG) models show great promise for large-scale screening and early detection of cardiovascular disease, even in low-resource settings.^8^ While the early results of these models are encouraging, clinical adoption remains limited.

A major obstacle to the widespread adoption of AI-ECG is the uncertainty surrounding its performance in real-world settings across diverse populations.^9^ To evaluate the generalizability of these models beyond their training population and to identify potential biases introduced by the training population, external validation is crucial.^10,11^ However, most AI-ECG models are not publicly available.^10^ As a result, external validation is often conducted by the original developers, raising concerns about biases like selective reporting and overfitting to specific populations.^10,12^

Among the various applications of AI-ECG, potentially one of the most clinically impactful and commonly studied is the detection of left ventricular systolic dysfunction (LVSD).^2,13,14^ Early and accurate detection of LVSD can significantly improve patient outcomes by enabling timely intervention and management.^15–17^ However, LVSD is often diagnosed at an advanced disease stage, which delays the timely initiation of therapy.^18^ Using AI-ECG models may provide a straightforward tool for screening patients at risk for LVSD who require further clinical evaluation.^19^

We sought to systematically compare the performance of AI-ECG models for LVSD detection in a standardized external cohort, while assessing the transparency and reproducibility of the field of AI-ECG. We identified all AI-ECG models through a systematic review and evaluated their internal and external performance, as well as the risk of bias of the identified models. We invited all authors to share their models for the first independent external validation and head-to-head comparison of AI-ECG for LVSD detection.

## Methods

The institutional review board (Medisch Etische Toetsingscommissie Amsterdam UMC) waived the requirement for informed consent since the study involved secondary analysis of existing data.

### Systematic review of AI-ECG for LVSD

To identify all available AI-ECG models for detecting LVSD, we conducted a systematic review of the literature. This systematic review was conducted according to the Preferred Reporting Items for Systematic Reviews and Meta-Analyses (PRISMA) guidelines **(Supplementary Table 1)**.^20^ Screening and data extraction were performed by two independent reviewers (PMC, MJB), and disagreements were resolved through discussion. The comprehensive methods for the systematic review are presented in the Supplementary Materials.

#### External validation cohort

Data for this study were obtained from a prospectively maintained registry that includes all consecutive patients who underwent cardiac magnetic resonance imaging (CMR) as part of routine clinical care, the Adam CMR database (Amsterdam UMC, The Netherlands). The database contains over 130 variables, including patient demographics, clinical characteristics, and detailed quantitative CMR-image measurements. All CMR scans were conducted between 2015 and 2022, and the reports were reviewed by expert cardiologists. From all patients in the Adam CMR database, clinically obtained 12-lead ECGs (Philips PageWriter or GE MUSE) were collected. All ECGs recorded within a 45-day window, covering 15 days before and 30 days after CMR, were included in the external validation cohort. LVEF was documented as a continuous variable for all patients and used to define the LVEF threshold labels.

To assess the models’ behavior in a lower complexity scenario, such as in the cardiology outpatient clinic or emergency department, we created a subcohort, referred to as the representative HF cohort. This cohort was constructed by stratified resampling from the full CMR cohort to include 15% of patients with LVEF <40% and 10% with LVEF between 40–50. While this cohort is a subset of the CMR cohort and not a real clinical cohort, the prevalence is in accordance with previously published reports of LVSD prevalence in these settings.^21,22^ Among the remaining controls, CMR findings were either normal or limited to isolated aortic pathology or coronary artery disease, limiting the number of complex cardiomyopathies with a normal LVEF. This stratification was performed to provide a sensitivity analysis to assess the models’ performances in a less complex setting and at the point of care where AI-ECG models are most likely to be applied, such as the emergency department or outpatient clinic. Additionally, since outcome prevalence can significantly impact model performance metrics, this design allows for the assessment of the influence of LVEF distribution on the discriminative performance. The use of CMR, known for its high precision in measuring LVEF, ensures robust ground truth labels, making it a particularly suitable modality for the AI-ECG evaluation.

#### External validation of included LVSD models

To gain access to existing AI-ECG models, we contacted all the corresponding authors of identified AI-ECG for LVSD publications. Initial emails were sent on July 16, 2024, outlining our request for independent external validation of their models. Specifically, we asked that the developed AI-ECG models for LVSD detection be shared for local deployment at Amsterdam UMC. If no response was received, a follow-up email was sent two weeks later to increase the likelihood of engagement and provide authors with a fair opportunity to share their models for independent validation. The email templates for the initial invitation and follow-up are provided in the Supplementary Methods. For all responding groups, meetings were held to discuss local technical deployment and concerns regarding data and model privacy. After these meetings, the research groups prepared their models for local deployment at Amsterdam UMC.

#### Model performance assessment

All shared models were applied to the ECGs in the CMR cohort to generate predicted probabilities for LVSD ranging from 0 to 1. The models were delivered as modules; either containerized using Docker or through repositories containing the entire pipeline and model weights. Thresholds were based on those provided by the original developers. Each module ingested raw signals since all signal-processing steps were integrated within the module. The number of ECGs for which models failed to produce valid predictions, for example, due to noise, was recorded and reported for each model. To ensure a fair comparison across models, sensitivity analyses were conducted using only the subset of ECGs that returned valid predictions for each model. Results were stratified by sex (male and female), age (<60 and ≥60 years), QRS duration (<100 ms and ≥100 ms), heart rate (>80 bpm and ≤80 bpm), atrial fibrillation status (present or absent), and clinical diagnosis related to the CMR, including coronary artery disease, ischemia, dilated cardiomyopathy, hypertrophic cardiomyopathy, myocarditis, and other cardiomyopathies. Additionally, false positive rates were calculated for LVEF 40-50% or LVEF >50%.

#### Sensitivity analyses

To determine if recalibrating the model thresholds could improve model performance in the Amsterdam UMC CMR dataset, we optimized the model probability threshold based on Youden’s Index within the Amsterdam UMC dataset. To assess the similarity of the outputs produced by the individual models, we calculated pairwise correlation coefficients using the predicted probabilities of each model.

#### Statistical analysis

The characteristics of the study population were summarized using means and standard deviations (SD) for continuous variables and counts and percentages for categorical variables. Model performance was evaluated by calculating the area under the receiver operating characteristic curve (AUROC), sensitivity, specificity, negative predictive value (NPV), and positive predictive value (PPV), along with 95% confidence intervals (CIs) derived from 500 bootstrap samples. For the representative HF cohort, a random distinct subset of the full cohort was used in each bootstrap iteration, fulfilling the pre-defined distributions. Calibration was evaluated by comparing predicted and observed risks across deciles of predicted probability. Pairwise correlations between model predictions were estimated using Spearman’s rank correlation coefficients. Analyses were conducted in Python 3.11.

## Results

### Identification and review of AI-ECG models

A systematic review of the literature identified 35 eligible studies describing 51 unique AI-ECG models for LVSD **(Table 1)**. The full results of the review are presented in the Supplementary Results, Supplementary Figure S1, and Supplementary Tables S4–S7. Most models were convolutional neural networks trained on ECG signal data, with LVSD mainly defined as LVEF <40% on echocardiography. Among these articles, 9 (25%) reported internal validation results, 11 (31%) included both internal and external validation, and 15 (43%) involved external validation only. All external validation efforts were carried out by the model developers.

**Table 1:**
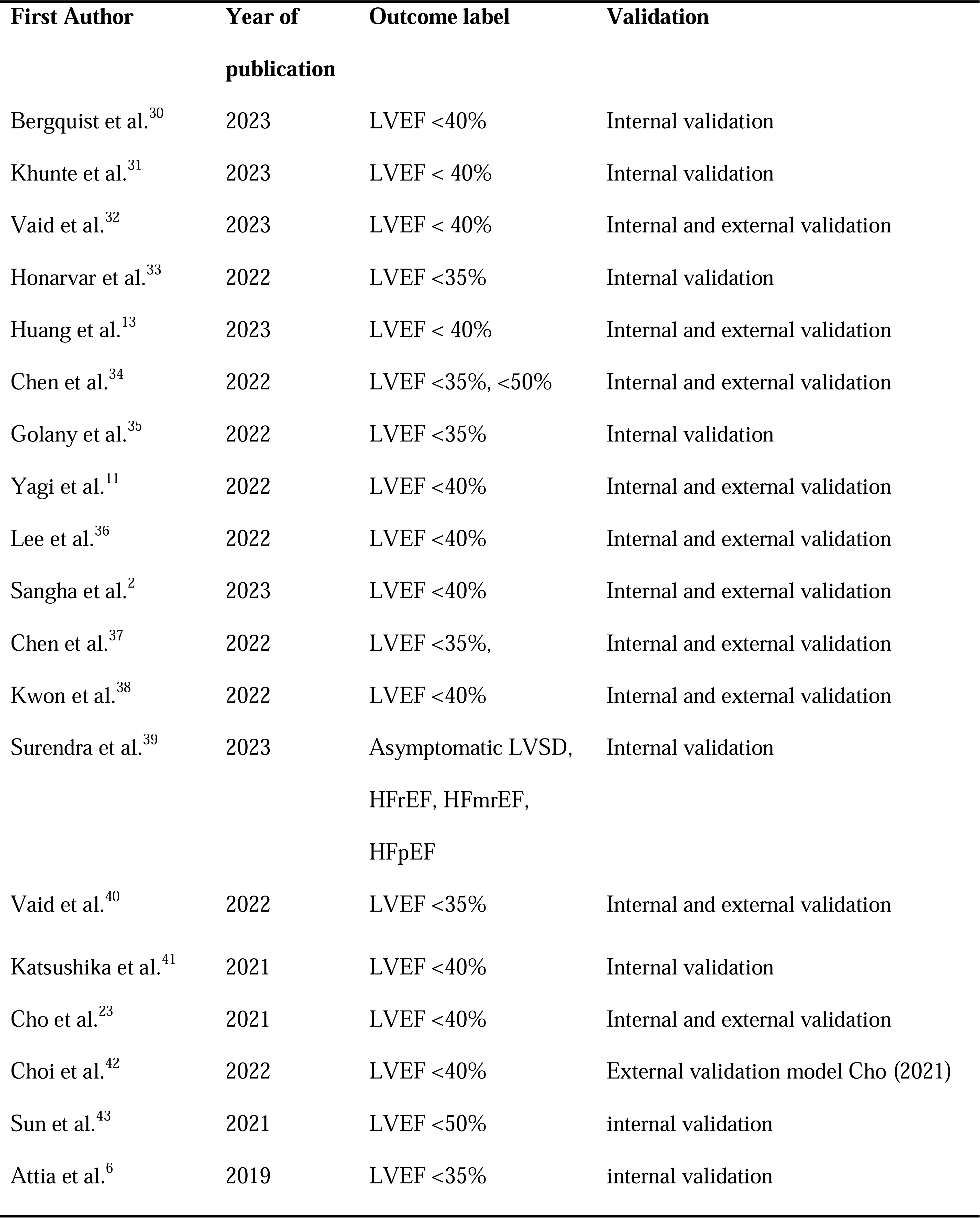
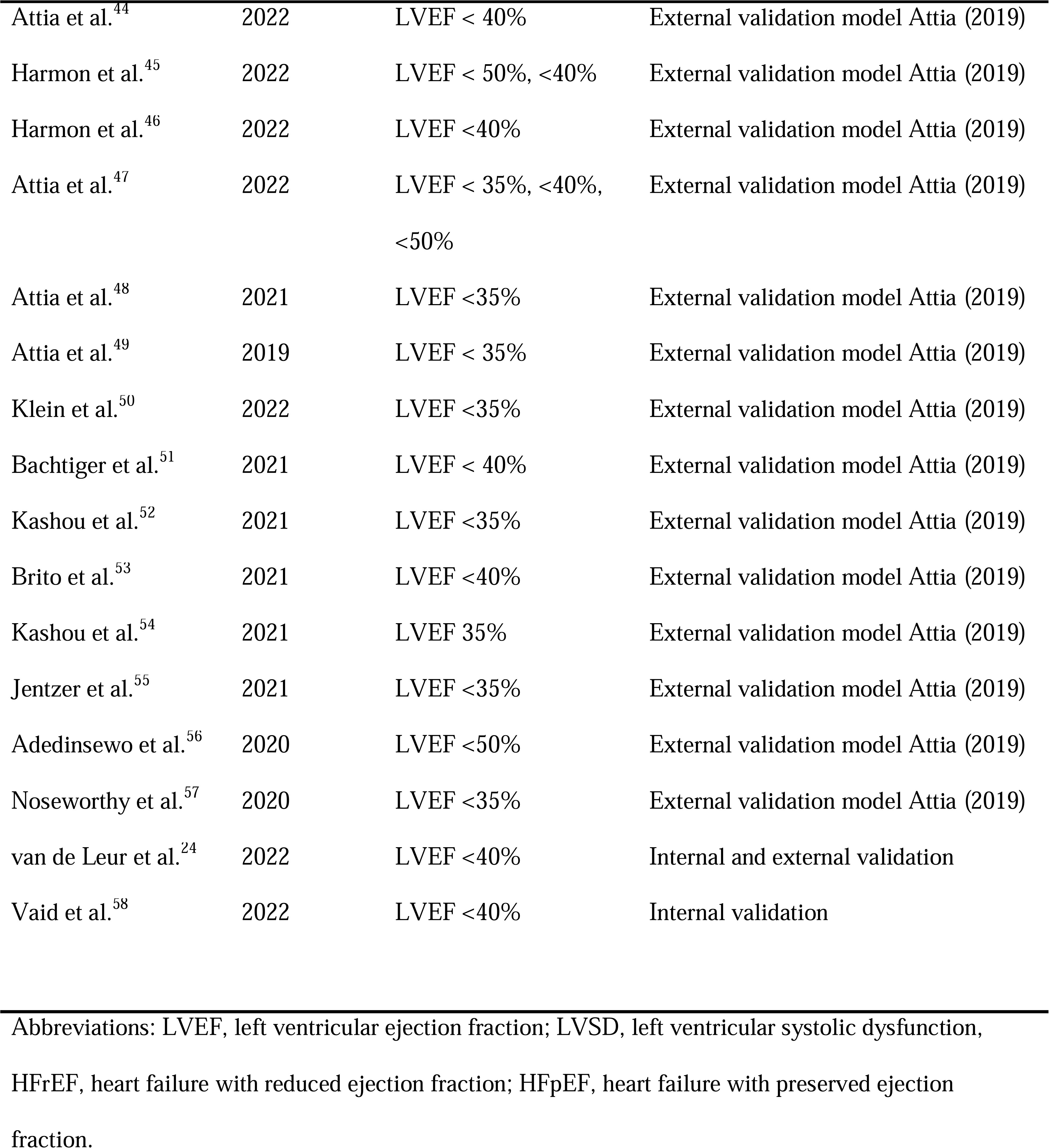
Overview of all included articles.

### Risk of bias of AI-ECG models

The description of baseline characteristics varied, with multiple studies not providing detailed cohort characteristics for both the development and external validation cohorts **(Figure 1A)**. In 12 of the 20 articles describing model derivation, external validation was reported either within the same article or in a separate article (Supplementary Table 6). For the internal validation, the performance was excellent in 29 (57%) of the models, good in 19 (37%), moderate in two (4%), and poor in one (2%), respectively **(Supplementary Table 6)**. For external validation, the performance was excellent in 31 (50%) of the models, good in 28 (45%), and moderate in three (5%), with none being poor **(Figure 1B, Supplementary Table 7)**. Of the articles reporting on the derivation of their models, nine stratified the results to identify potential bias toward subgroups based on age, sex, ethnicity, comorbidity, or ECG characteristics **(Supplementary Table 6)**.

**Figure 1:**
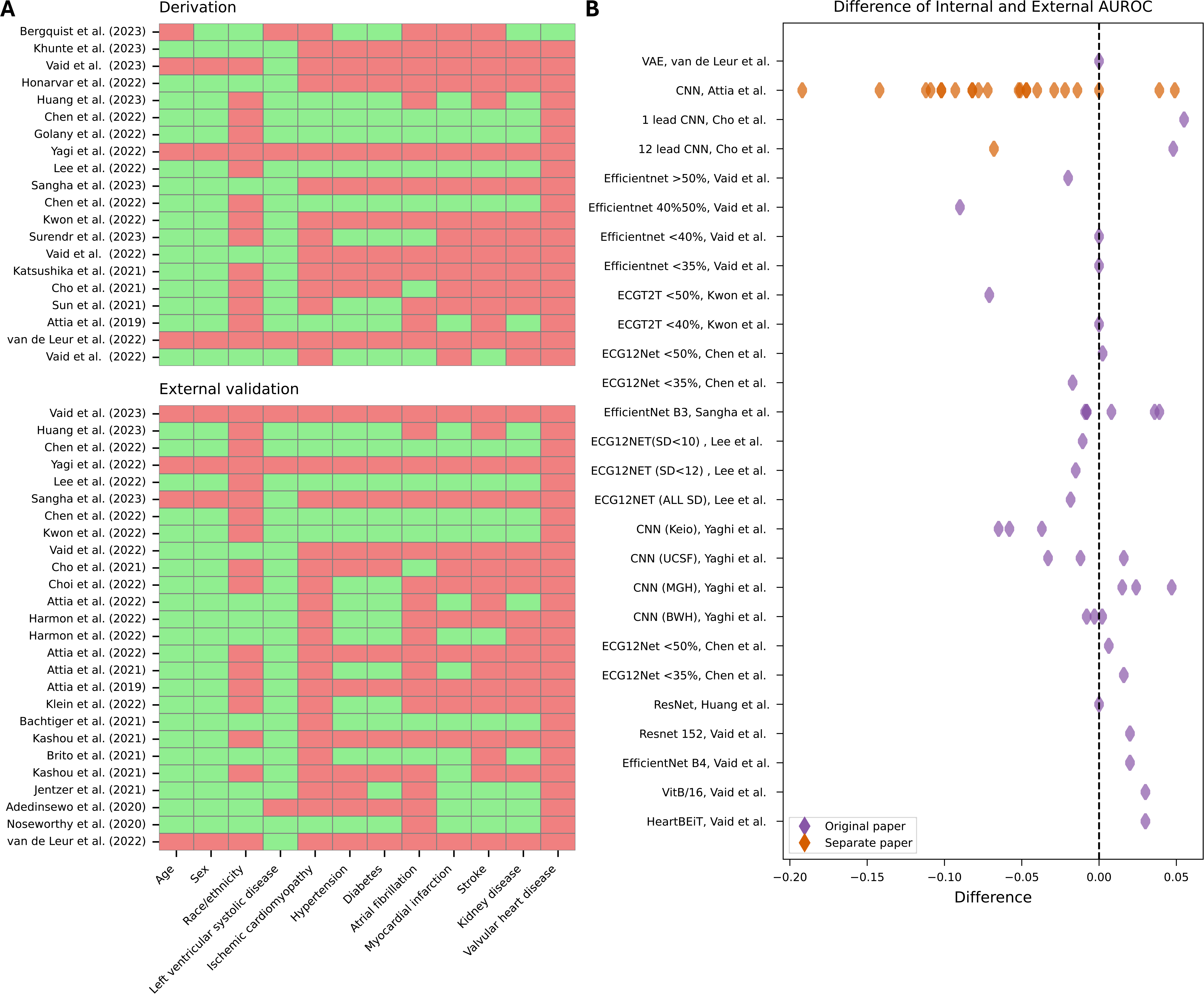
Reported baseline characteristics, internal and external performance for included models. Panel A: These heatmaps display baseline characteristics reported in derivation (top) and external validation (bottom) studies. Each row represents a study, and each column indicates a specific characteristic (e.g., age, sex, comorbidities). Green shows that the characteristic was reported, while red indicates it was not. Panel B: The difference between internal validation AUROC and external validation AUROC across models. The purple diamonds represent external validation reported in the original paper. The red diamonds show external validation conducted in separate papers. All values are referenced to internal AUROC; therefore, a value higher than zero indicates a higher internal AUROC compared to the AUROC during external validation. Abbreviations: AUROC, Area Under the Receiver Operating Curve; CMR, Cardiac Magnetic Resonance; HF, Heart failure.

According to the PROBAST assessment, the majority of derivation studies were rated as having high or unclear risk of bias, mainly due to a lack of external validation or inadequate cohort descriptions **(Supplementary Figures 2 and 3)**. Concerns about applicability were infrequent **(Supplementary Figures S2–S3)**.

### Model availability

None of the reported models and/or weights were publicly available for independent external validation. After contacting all corresponding authors of articles describing the development of AI-ECG models for LVSD detection, six research groups responded to our inquiries. Of these, four research groups agreed to participate in the external validation study by sharing their models for external validation. The models originated from Korea, the United States (US), Taiwan, and the Netherlands.

The research group identified through Cho et al. shared an algorithm called AiTiALVSD v2.00.00, which is based on a transformer architecture and convolutional encoders.^23^ It was trained on 498,726 ECGs from 186,889 patients across 16 tertiary hospitals in Korea, using an 80–10–10% split. Labels were defined as LVEF <40% on TTE measurement within 30 days of the included ECG. External validation performance of this version of the model has not yet been published.

Sangha et al. introduced an AI-ECG model called ECG Vision.^2^ This model utilized a pre-trained version of EfficientNet-B3, which was fine-tuned using a dataset comprising 116,210 patients from Yale New Haven Hospital in the US, with an 85-5-10% split for training, validation, and testing. The model inputs are 300×300 ECG images, either directly or plotted from raw signals. LVSD was defined as LVEF <40% based on TTE measurements performed within 15 days of the ECG. The model achieved an AUROC of 0.90–0.95 across six external validation sites (Supplementary Tables 6 and 7).

Huang et al. shared a ResNet-based model trained on signal data from 380,675 patients at Chang Gung Memorial Hospital (CGMH) in Taiwan, using a 35-15-50% split for training, validation, and testing, which will be referred to as the CGMH model.^13^ LVSD was defined as LVEF <40% on TTE performed within two weeks of the ECG. External validation was performed on data from another hospital in Taiwan, resulting in an AUROC of 0.94 for internal validation and 0.95 for external validation **(Supplementary Tables 6 and 7)**.

The research group, as identified through the publication by van de Leur et al.^24^, shared an unpublished ResNet-based model trained on median beat ECGs from 57,250 patients across two hospitals in the Netherlands. They used a 90-10% split for training and internal validation and evaluated it on a different internal cohort of newly referred cardiology outpatients. This model differs from the one in the original publication, with differences in architecture, training, and development cohort. The pipeline applied a custom algorithm to construct median-beat ECGs. As median-beat reconstruction requires consistent and noise-free beats, this preprocessing step was more sensitive to signal noise, leading to rejection of 6% of ECGs. LVSD was defined as LVEF <40% on TTEs performed within 90 days of the ECG.

### External validation of AI ECG models for LVSD detection

Independent external validation was conducted using the full CMR cohort, which included 1,203 patients and 4,737 corresponding ECGs, with one randomly selected ECG per individual in each bootstrap iteration. The average age of the cohort was 59 ± 15 years, and 431 (36%) were female. Based on CMR measurements, left ventricular ejection fraction (LVEF) was <40% in 212 (18%) patients, between 40% and 50% in 231 (19%) patients, and >50% in the remaining 760 (63%) patients **(Table 2).** The subset of patients for which all models provided valid predictions included 94% of all ECGs. For the representative heart failure (HF) cohort, 554 patients were included in each bootstrap iteration, consisting of 84 (15%) with LVEF <40%, 50 (9%) with LVEF between 40–50%, and the remaining 420 (76%) with LVEF >50%.

**Table 2:** Baseline characteristics of the Amsterdam UMC CMR-cohort.

| Characteristic | n (%) or mean ± SD |
| --- | --- |
| Number of ECGs | 4737 |
| Number of unique Patients | 1203 |
| Age | 59.5 ± 14.9 |
| Female | 431 (35.9) |
| LVEF 40-50 | 231 (19.3) |
| LVEF<40 | 212 (17.7) |
| Coronary artery disease | 328 (27.6) |
| Cardiomyopathy other | 147 (12.4) |
| Ischemia | 87 (7.3) |
| Dilated cardiomyopathy | 83 (7.0) |
| Aortic pathology | 49 (4.1) |
| Hypertrophic cardiomyopathy | 63 (5.3) |

2 Diagnoses are based on the conclusions of the cardiac magnetic resonance imaging study reported 3 by an expert cardiologist. Abbreviations: n, number of samples; SD, Standard Deviation; LVEF, 4 Left Ventricular Ejection Fraction.

### Performance of AI-ECG Models in Detecting Left Ventricular Dysfunction

AiTiALVSD achieved an AUROC of 0.94 (CI: 0.93;0.94) in the full CMR cohort and 0.96 (CI: 0.94;0.97) in the representative HF cohort **(Figure 2A, Supplementary Tables 8-9).** ECG Vision achieved an AUROC of 0.89 (CI: 0.88;0.90) in the full CMR cohort and 0.92 (CI: 0.89;0.94) in the representative HF cohort. The AUROC for CGMH was 0.90 (CI:0.89;0.91) in the full CMR cohort and 0.94 (CI:0.92;0.95) in the representative HF cohort, while the Utrecht model achieved an AUROC of 0.83 (CI: 0.82;0.84) and 0.86 (CI: 0.83;0.89) in these cohorts, respectively. Sensitivity analyses limited to ECGs with valid predictions across all models demonstrated no meaningful change in model performance **(Supplementary Table 10)**.

**Figure 2:**
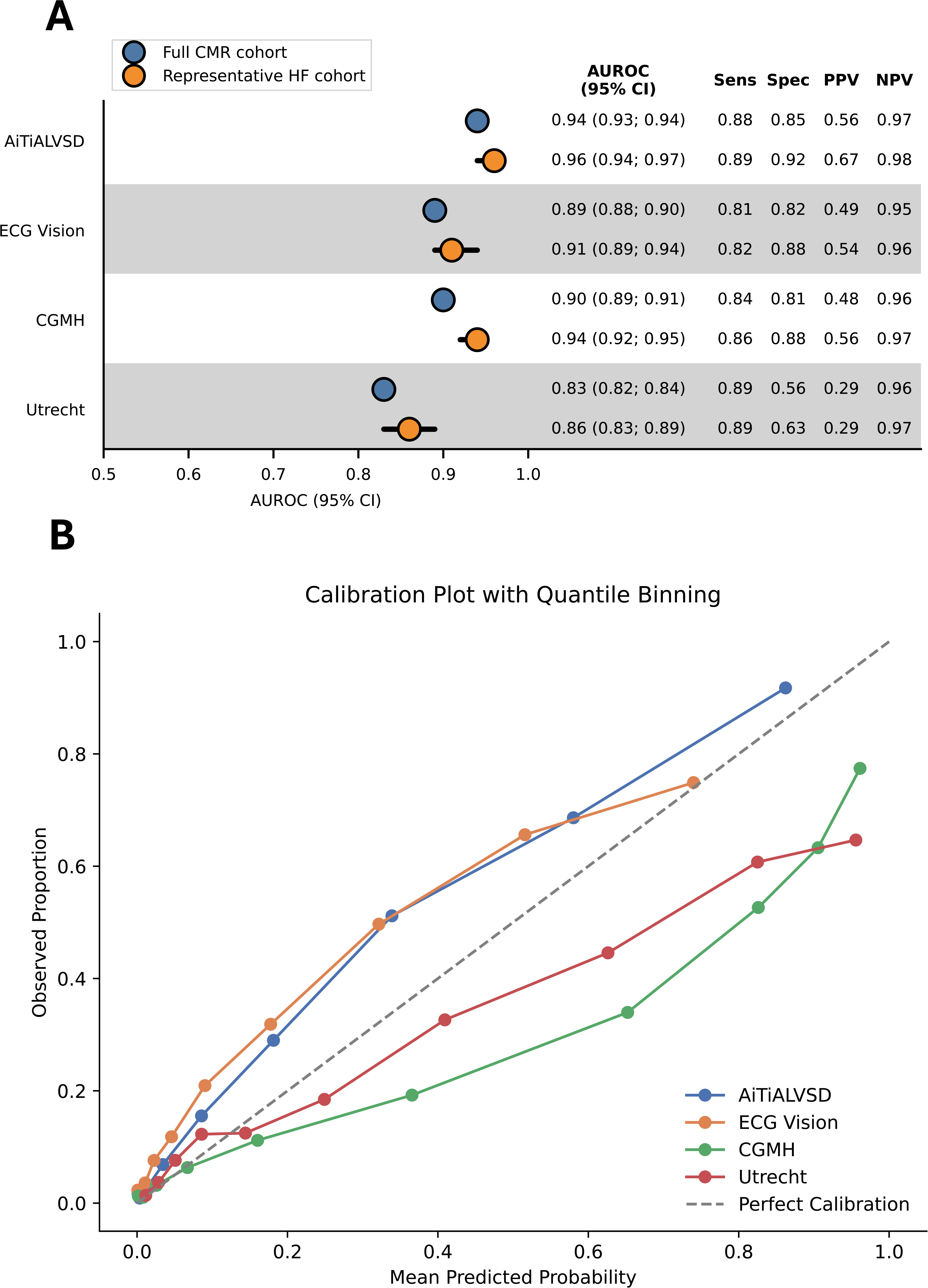
Performance and calibration of included models for external validation. Panel A: Forest plot showing the independent external validation results of four models (Cho et al., Sangha et al., Huang et al., and van de Leur et al.) in the full CMR cohort and the augmented heart failure cohort. The plot displays AUROC values with their 95% confidence intervals (CI). Panel B: Calibration plot for the external validation. Mean predicted probabilities versus observed proportions, shown using quantile binning (10 bins). The “Perfect Calibration” line indicates ideal model calibration. Abbreviations: CMR, Cardiac Magnetic Resonance imaging; HF, Heart Failure; AUROC, Area Under the Receiver Operating Characteristic Curve; Sens, Sensitivity; Spec, Specificity; PPV, Positive Predictive Value; NPV, Negative Predictive Value.

The AiTiALVSD and ECG Vision slightly underestimated the observed risk, while the models CGMH and Utrecht slightly overestimated it **(Figure 2B)**. Recalibrating the thresholds caused small differences in sensitivity and specificity. In three models, sensitivity improved slightly at the cost of a minor decrease in specificity, whereas in one model, specificity increased slightly with a corresponding drop in sensitivity **(Supplementary Table 11)**.

Stratification by age and sex did not reveal any potential bias **(Figure 3, Supplementary Tables 8 and 9)**. All four models demonstrated slightly reduced performance in ECGs with a QRS width greater than 100ms and in patients with atrial fibrillation. Additionally, AUROCs were generally lower in subgroups with cardiomyopathy. Despite these differences, the models demonstrated good to excellent performance across all subgroups **(Figure 3)**. The false positive rates ranged from 53% to 65% in patients with an LVEF between 40% and 50%, and from 12% to 39% in patients with an LVEF greater than 50% for the included models **(Supplementary Table 12)**.

**Figure 3:**
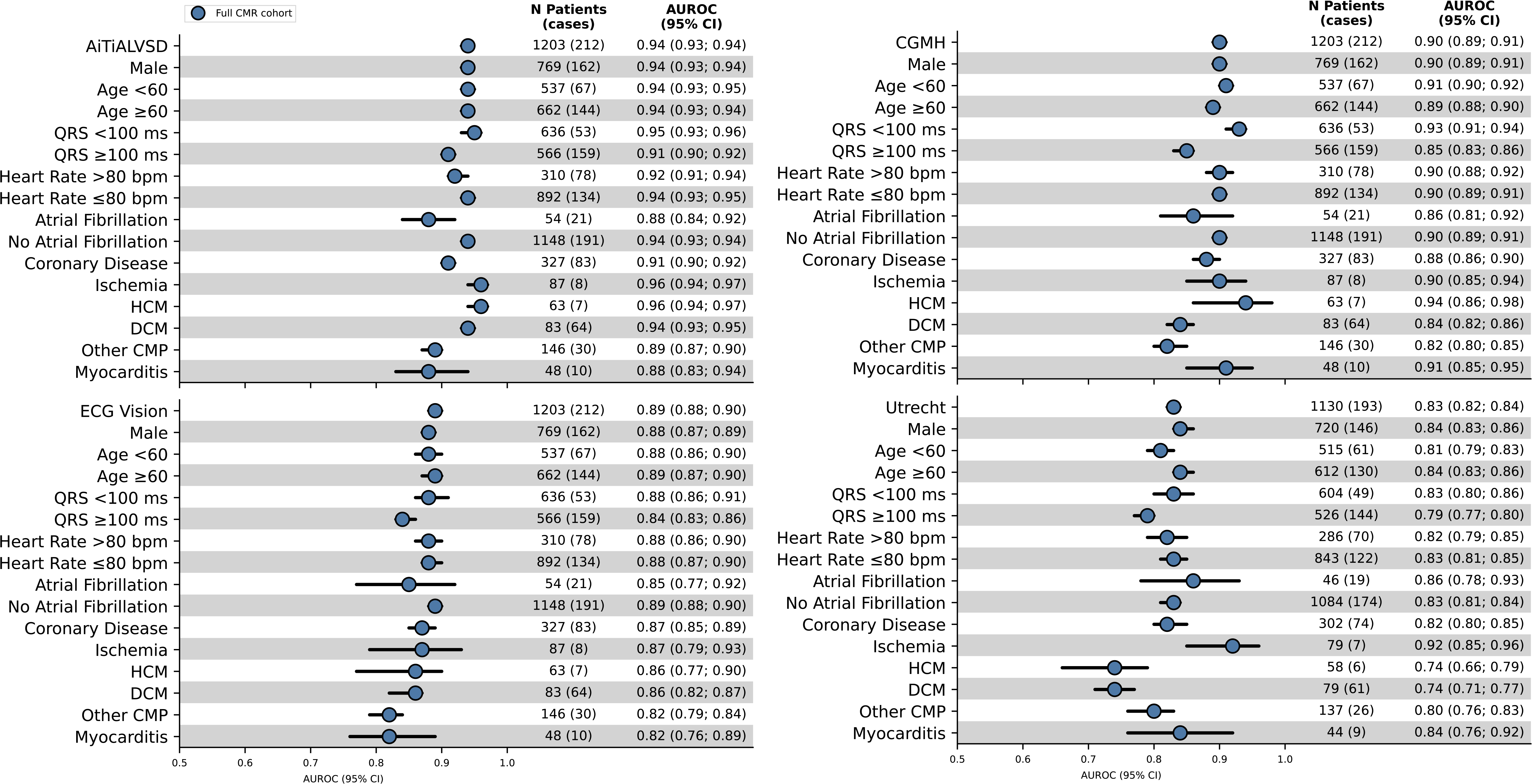
Subgroup analysis of AUROC values for external validation of models. Performance in the full CMR cohort across various patient subgroups is shown, including sex, age, QRS duration, heart rate, atrial fibrillation status, coronary disease, types of cardiomyopathies, and myocarditis. The total number of patients is listed, along with the number of cases with LVSD in the chosen category (n patients (cases EF<40%)). CMR; Cardiac Magnetic Resonance imaging, AUROC values are provided with 95% confidence intervals to give insights into model performance within specific populations. Abbreviations: AUROC, Area Under the Receiver Operating Characteristic Curve; HCM, Hypertrophic Cardiomyopathy; DCM, Dilated Cardiomyopathy; CMP, Cardiomyopathy.

### Correlation Between Individual Models

Pairwise Spearman’s correlation coefficients of the continuous model outputs demonstrated moderate to high correlations across all models **(Figure 4)**. The highest correlation was observed between the CGMH model and AiTiALVSD (ρ = 0.86). Correlations with the Utrecht model were generally lower (ρ 0.69–0.73).

**Figure 4:**
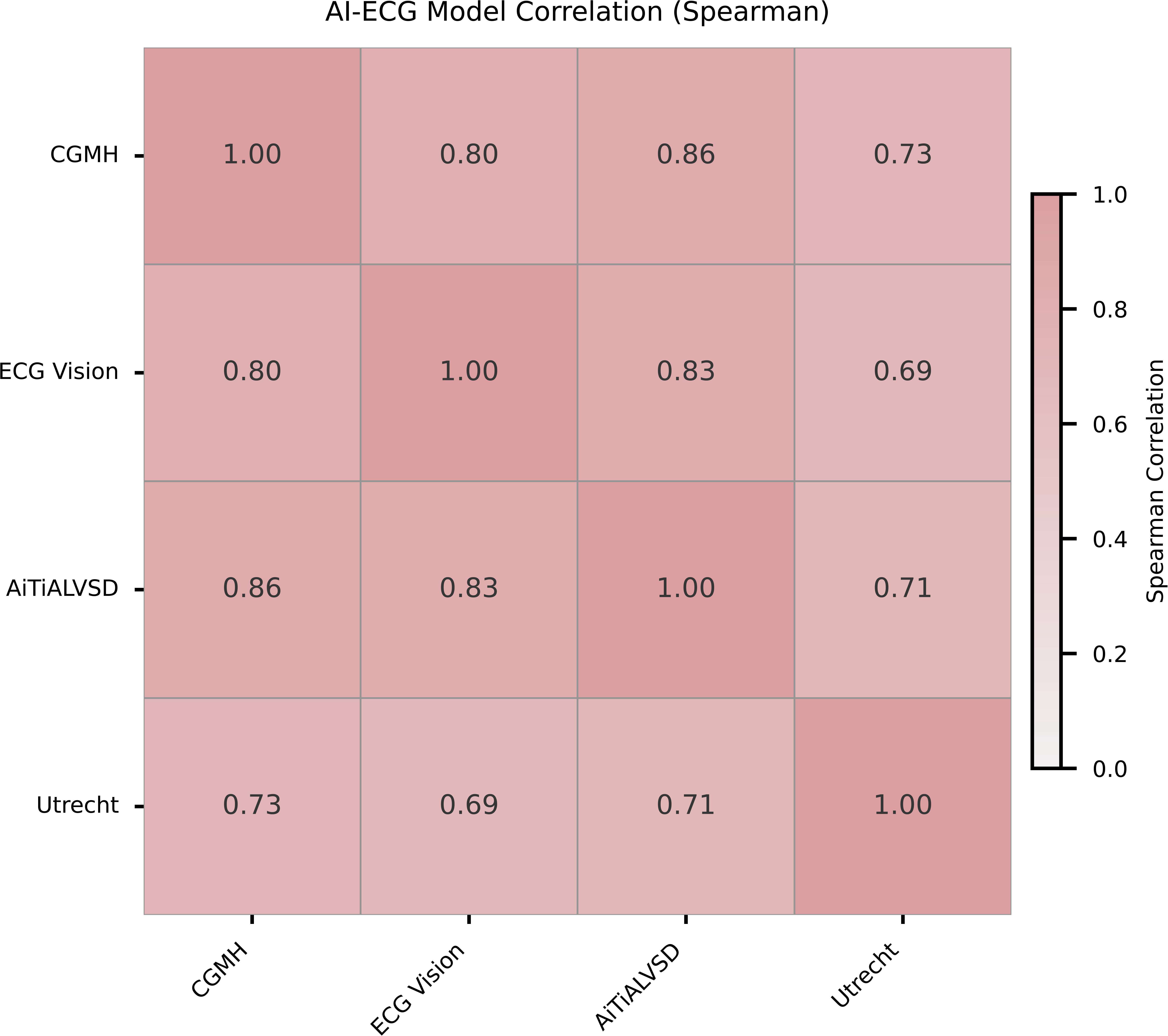
Correlation between the different AI-ECG models. The heatmap shows the pairwise Spearman correlation coefficients between the model predictions of the different included models. Abbreviations: CMR, Cardiac Magnetic Resonance imaging; HF, Heart Failure; AUROC, Area Under the Receiver Operating Characteristic Curve; Sens, Sensitivity; Spec, Specificity; PPV, Positive Predictive Value; NPV, Negative Predictive Value.

## Discussion

We are the first to introduce a new framework for the external validation of AI-ECG, providing a systematic, head-to-head comparison of all available AI-ECG models for LVSD detection. We found that the overall risk of bias for AI-ECG remains high, mainly due to incomplete descriptions of training cohorts and an overreliance on developer-led validations. Importantly, of the 51 models we identified, only four were accessible for independent testing, highlighting significant gaps in transparency and raising important questions about external validity. The four models available for external validation on a single, well-characterized dataset performed consistently despite differences in training populations from different continents. Our research shows that their diagnostic accuracy depends on edge-case examples, notably individuals with an LVEF between 40 and 50%, where false positives are common. High inter-model correlations indicate that AI-ECG models are capturing similar electrocardiographic signs of LVSD. Furthermore, validation in a more complex cohort attenuates the diagnostic discrimination ability, underscoring the importance of using representative validation populations.

The strong performance of AI-ECG models for LVSD detection in the literature underscores their potential to screen patients for early diagnosis of LVSD. However, this study reveals significant gaps in the evaluation of AI-ECG models, particularly in assessing potential biases, which may undermine their trustworthiness and hinder clinical adoption. External validation in all reviewed articles was performed solely by the original developers, increasing the risk of selective data inclusion and biased reporting. Moreover, most models do not fully align with the FUTURE-AI guidelines, particularly in terms of transparency, reproducibility, and open access.^10^ With the recent FDA approval of AI-ECG models, which signals their potential integration into clinical practice, it is crucial to evaluate the models’ performance while minimizing potential bias. This makes it even more concerning that only a small number of AI-ECG models were made available for our external validation study. Selective sharing of models likely introduces systematic bias, as investigators who are more confident in their models may be more willing to share them, creating an inflated and potentially misleading perception of performance at the field level.

The four models shared for independent external validation demonstrated consistent performance, with a slight attenuation of the AUROC when comparing the lower-complexity (representative HF) cohort to the higher-complexity (full CMR) cohort. Notably, this external performance remained consistent, even though three of the models were trained on geographically distinct cohorts (Korea, the US, and Taiwan). Moreover, the continuous probabilities produced by these models exhibited high pairwise correlation, indicating that AI-identified ECG features are consistent despite being trained on three continents, which supports their generalizability. Additionally, models trained on raw ECG signals or ECG images performed similarly well, aligning with recent literature findings.^8,25–27^ It is essential to note that all included AI-ECG models were initially developed and validated using TTE-derived LVEF as the ground truth. In contrast, our independent validation utilized CMR, the reference standard for LVEF assessment, known for its superior accuracy and reproducibility. Despite the differences between these modalities, with TTE tending to underestimate LVEF compared to CMR, the models maintained strong performance, highlighting their robustness and potential clinical utility. The slight performance decline observed in the full CMR cohort warrants careful consideration and possibly model recalibration when implemented in highly complex patient populations.

Our findings demonstrate that the characteristics of the validation cohort substantially influence model performance. An AUROC difference of ∼0.04 between the low-complexity (representative HF) and high-complexity (CMR referral) populations underscores this effect. Many published models fail to clearly describe their cohort composition, which limits their interpretability and applicability.^28^ Performance variation across subgroups, such as those with broad QRS or atrial fibrillation, further highlights the need for transparent reporting and potential model recalibration.^29^ Notably, most false positives in our study had CMR-derived LVEF between 40–50%, suggesting borderline dysfunction and reflecting the challenge of distinguishing subclinical or overlapping cardiac conditions.

### Limitations and Future Directions

We included four models for external validation, all of which showed good or excellent performance in our external validation. However, this might have introduced publication bias, as research groups with better-performing models may have been more willing to share them. Additionally, some of the research groups in the external validation provided updated versions of their models that differ from those in the systematic review, precluding direct comparison of their internal and external performance. Furthermore, even though all models were trained to detect LVSD defined as LVEF <40%, heterogeneity in the input format of the ECG and development pipelines limits direct equivalence of performance estimates. Another important limitation is that the analysis was limited to a single Western European cohort of patients referred for CMR, representing a more challenging classification task and enrichment of complex cardiomyopathies. To mitigate the effect of cohort-specific selection bias, we simulated a population with a prevalence similar to that in the emergency departments or outpatient clinics. This analysis should be interpreted as a sensitivity analysis since it is an augmented cohort, and not a real clinical cohort. Finally, we used CMR-derived LVSD to define the outcome of interest. While CMR is considered the reference standard for LVEF assessment, it systematically differs from TTE-derived LVEF, which was used during model development. While CMR allows for a more accurate evaluation, this modality mismatch may have influenced model performance, particularly in patients with borderline LVEF values.

Future research should prioritize collaborative efforts, sharing models, and independent external validation across datasets that are geographically and demographically diverse. Additionally, using standardized metrics and benchmarks to describe dataset features will be crucial for ensuring consistency and comparability in the literature. With these combined efforts, AI-ECG models can evolve from promising research tools to actual clinical decision-support systems, providing real benefits to patients.

## Conclusion

AI-ECG models for detecting LVSD demonstrated high discriminative performance in a novel framework for independent external validation. However, the incomplete reporting of development and validation cohort characteristics in many studies raises concerns about potential bias, and the limited availability of models greatly hinders independent validation, thereby impeding wider clinical adoption.

## Author contributions

PMC and MJB designed the study. PMC and MJB performed the study selection and data extraction and drafted the manuscript. PMC performed the analyses and implemented the models. MMW, FWA, CPA, JLS provided critical input on the analysis, as well as the drafted manuscript. BA, LSD, YH, TM, RK, CK, JK, HL, MSL, RRL, ZL, EKO provided their models for external validation and reviewed the final draft of the manuscript for correctness.

## Data availability

Due to the inclusion of patient-level data, the dataset cannot be publicly shared; however, researchers may contact the corresponding author to request access for academic purposes.

AI-ECG: Artificial intelligence–enhanced electrocardiogram
LVSD: Left ventricular systolic dysfunction
LVEF: Left ventricular ejection fraction
CMR: Cardiac magnetic resonance
TTE: Transthoracic echocardiography
PROBAST: Prediction model Risk Of Bias ASsessment Tool
PRISMA: Preferred Reporting Items for Systematic Reviews and Meta-Analyses
CGMH: Chang Gung Memorial Hospital

## Central Illustration

We propose a structured framework for the independent validation of artificial intelligence–enabled electrocardiography (AI-ECG) models. Of 51 published AI-ECG models for left ventricular systolic dysfunction (LVSD), only four were available for true external validation. These models were evaluated in an independent cohort with paired electrocardiograms and cardiac magnetic resonance imaging (CMR) as the reference standard. Despite differences in architecture and training cohorts, model performance remained high in this external setting. However, limited openness and accessibility of AI-ECG models constrain independent assessment and may impede broader clinical adoption.

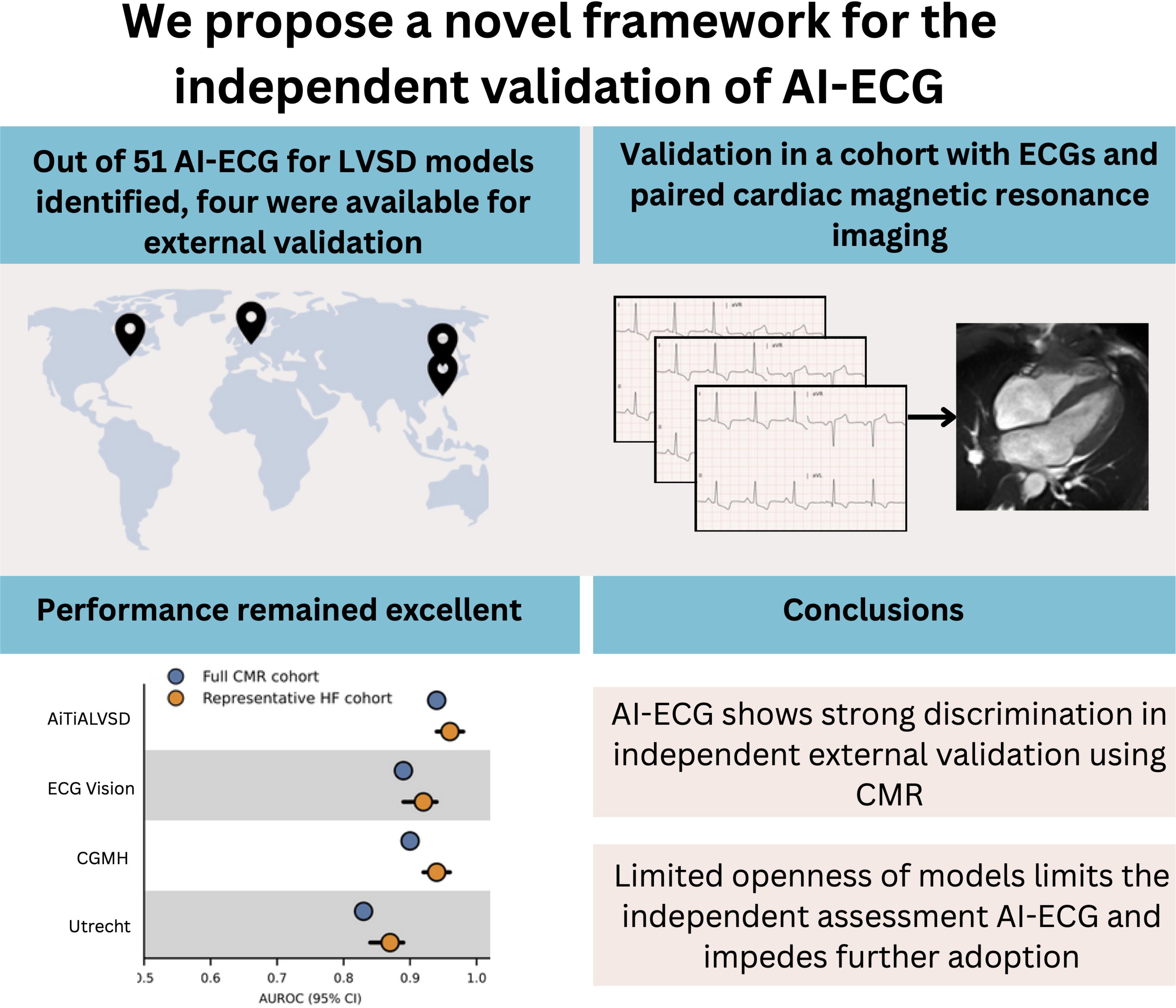

