## Supplementary Materials for "Artificial Intelligence-Enhanced Electrocardiogram Models for Detection of Left Ventricular Dysfunction: A Comparison Study"

**AI-ECG for LVSD detection: a first-in-kind head-to-head comparison of multinational models**

Croon et al.

*Supplementary Materials*

**Index**

| Supplementary Methods | p. 2 |
| --- | --- |
| Supplementary Figures | p. 4 |
| Supplementary Tables | p. 6 |

**Supplementary Methods**

This systematic review was conducted according to the Preferred Reporting Items for Systematic Reviews and Meta-Analyses (PRISMA) guidelines (Supplementary Table 1). The institutional review board waived the requirement for informed consent since the study involved secondary analysis of existing data.

### *Search Strategy*

The Ovid MEDLINE(R) electronic database was systematically searched in October 2023 to identify articles published since January 2010. The search strategy was designed to identify studies reporting on the development and/or external validation of AI-ECG models for the detection of LVSD (Supplementary Table 2). *Search Strategy*

A comprehensive search of the Ovid MEDLINE(R) database was conducted on October 5, 2023. The search strategy included terms related to electrocardiography (e.g., ECG, EKG, SAECG, "12-Lead ECG") and various machine learning methodologies (e.g., deep learning, convolutional neural networks, autoencoders, supervised learning, generative adversarial networks). Identified studies were imported into Rayyan for screening and deduplication. The search was limited to studies involving human subjects, published from 2010 onwards. Animal studies, case reports, reviews, editorials, and comments were excluded. The employed search strategy is provided in Supplementary Table 1.

*Exclusion criteria*

Articles were excluded based on the following criteria:
- Deep learning applied to manually derived ECG features
- Focus on exercise ECG
- Long-term ECG recordings (>30 seconds) or Holter monitoring
- Natural language processing applied to ECG reports
- Cardiac age predictors
- Image-based source data
- Heart rate variability analysis
- Signal decomposition
- ECG filtering or data augmentation techniques
- Beat delineation and detection (e.g., onset-offset annotation)
- Pediatric studies (age <18 years)
- Animal studies
- Biometrics or authentication techniques
- Cost-effectiveness analyses of AI-ECG
- Non-standard ECG devices (e.g., defibrillators or implanted devices)
- Photoplethysmography (PPG) analysis
- Conference papers
- Data Collection and Analysis

### *Study selection criteria*

All articles describing the development and/or external validation of deep learning (DL) models using ECG data, hereafter referred to as AI-ECG models, were considered for inclusion. We included both standard 12-lead ECGs and reduced-lead recordings (e.g., single-lead wearable data). Studies analyzing ECG signals exceeding 30 seconds in duration were excluded to differentiate AI-ECG models from those applied to continuous ECG recordings, such as Holter or telemetry recordings. From the eligible studies, we only selected those specifically focused on the development and/or external validation of AI-ECG models for detecting LVSD. A detailed overview of the exclusion criteria is provided in the Supplementary Methods.

*Screening process*

All articles were screened for eligibility by two independent reviewers (PC and MB) based on predefined study selection criteria using titles and abstracts. Disagreements were resolved through discussion. Articles that appeared potentially relevant were retrieved in full-text and subsequently reassessed for eligibility by the same reviewers.

### *Data synthesis and analysis*

Both reviewers independently extracted data using a standardized data extraction form to ensure the completeness and accuracy of the data. General characteristics of the developed models, including their architecture, training cohort, internal and external validation cohort characteristics, data and model availability, and model performance metrics, were extracted. A complete list of the extracted variables is provided in Supplementary Table 3. Continuous variables are reported as mean ± standard deviation or median with interquartile range, and categorical variables are presented as counts with percentages. The performance of models was categorized into four groups: excellent (area under the receiver operating characteristic curve [AUROC] > 0.9), good (0.8-0.9), moderate (0.7-0.8), and poor (<0.7).

*Email template initial invitation*

Dear [NAME CORRESPONDING AUTHOR]

I hope this message finds you well. My name is Philip Croon, and I am currently conducting a systematic review of AI ECG models for detecting left ventricular dysfunction under the supervision of Prof. Folkert Asselbergs. We are reaching out regarding your published LVEF deep learning model described in your paper titled "[NAME OF ARTICLE]".

As part of our review, we aim to perform a 'head-to-head' comparison of the included models by externally validating all available AI-ECG models using a clinical database from Amsterdam UMC. We are keen to locally implement your model to assess its generalizability and performance with our internal dataset.

Would it be possible to receive your model for this purpose? We believe this step is crucial for advancing the field and ensuring the robustness of AI-driven diagnostics.

Thank you for considering our request. We look forward to your positive response and the possibility of collaborating.

Best regards,

*Email template follow-up*

Dear [NAME CORRESPONDING AUTHOR],

I hope this message finds you well.

I am writing to follow up on my email sent on July 16th regarding a potential collaboration for a review project. We are conducting a head-to-head comparison of deep learning ECG models for detecting left ventricular dysfunction and would like to externally validate your AI Model.

We would greatly appreciate your consideration of our invitation to collaborate on this review.

Thank you for your time and consideration. I look forward to your response.

Best regards,

**Supplementary results**

A total of 4,321 articles were identified in the search. After removing 70 duplicate records, 4,251 articles remained for title and abstract screening, during which 3,680 articles were excluded **(Supplementary Figure 1)**. An additional 188 articles were excluded during full-text screening. The most common reasons for exclusion were the use of ECG recordings > 30 seconds (e.g., Holter monitoring or telemetry), beat detection models, and manual ECG-feature extraction. Ultimately, only articles describing the derivation and/or validation of AI-ECG for LVSD were included in the analysis. Therefore, all articles (n=348) that were not primarily trained for LVSD, did not describe derivation and/or validation, used multi-modal data, or lacked a clear outcome definition were excluded, resulting in 35 articles included in the final analysis **(Table 1)**.

Of the included articles, 9 (25%) reported internal validation performance only, 11 (31%) both internal and external validation, and 15 (43%) external validation only. All external validation efforts were conducted by the model developers. Of the included articles describing the development of a model, 10 (50%) originated from Asia, 8 (40%) from North America, and 2 (10%) from Europe. In total, all articles included 51 different AI-ECG models for the prediction of LVSD. All models were variations of convolutional neural networks. Of these models, 48 (94%) utilized direct signal data, and 3 (6%) used an ECG-image as input for the model **(Supplementary Tables 4-7)**.

### *LVSD definitions*

In 34 articles covering 50 models, LVSD was defined using LVEF measured via transthoracic echocardiography (TTE). The cutoff to define LVSD was set at LVEF <35% for nine models, LVEF <40% for 31 models, LVEF between 40% and 50% in two models, and LVEF <50% in six models. One article did not specify a TTE-based LVEF cutoff for defining LVSD. Instead, it used disease definitions such as HF with reduced EF, HF with mildly reduced EF, HF with preserved EF, and asymptomatic LVSD.

*Quality and bias assessment*

According to the PROBAST checklist, among all studies reporting on the derivation of AI-ECG models, eight models (40%) were rated as having a high risk of bias, and another eight models (40%) had an unclear risk of bias (Supplementary Figure 2). Concerns regarding applicability were high in 1 model (5%) and unclear in 2 models (10%) (Supplementary Figure 3). High risk of bias was exclusively due to the lack of external validation. The unclear risk of bias primarily resulted from insufficient or vague descriptions of the study cohort or cohort characteristics, which did not align with the intended real-world applications (e.g., unusually low outcome prevalence). High or unclear concerns about model applicability arose from an undefined or poorly defined outcome (n = 1, 5%), outcome definitions that did not reflect real-world clinical scenarios (n = 1, 5%), or participant inclusion criteria that conflicted with typical clinical practice (n = 1, 5%).

**Supplementary Figures**

**Supplementary Figure 1** Flowchart of the Study Selection of article’s describing the development or validation of AI-ECG Models for LVSD Detection

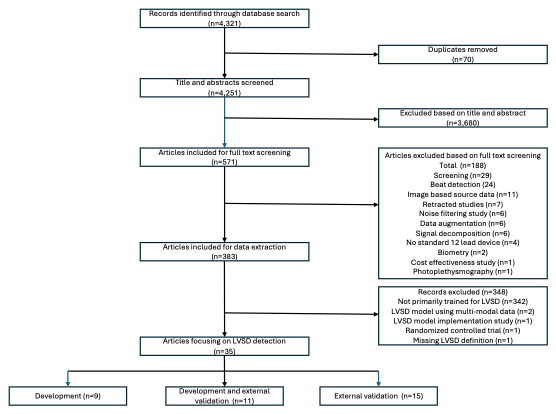

**Abbreviations: LVSD, Left Ventricular Systolic Dysfunction.**

**Supplementary Figure 2.** Summary of risk of bias assessment according to PROBAST (Prediction model Risk Of Bias ASsessment Tool)

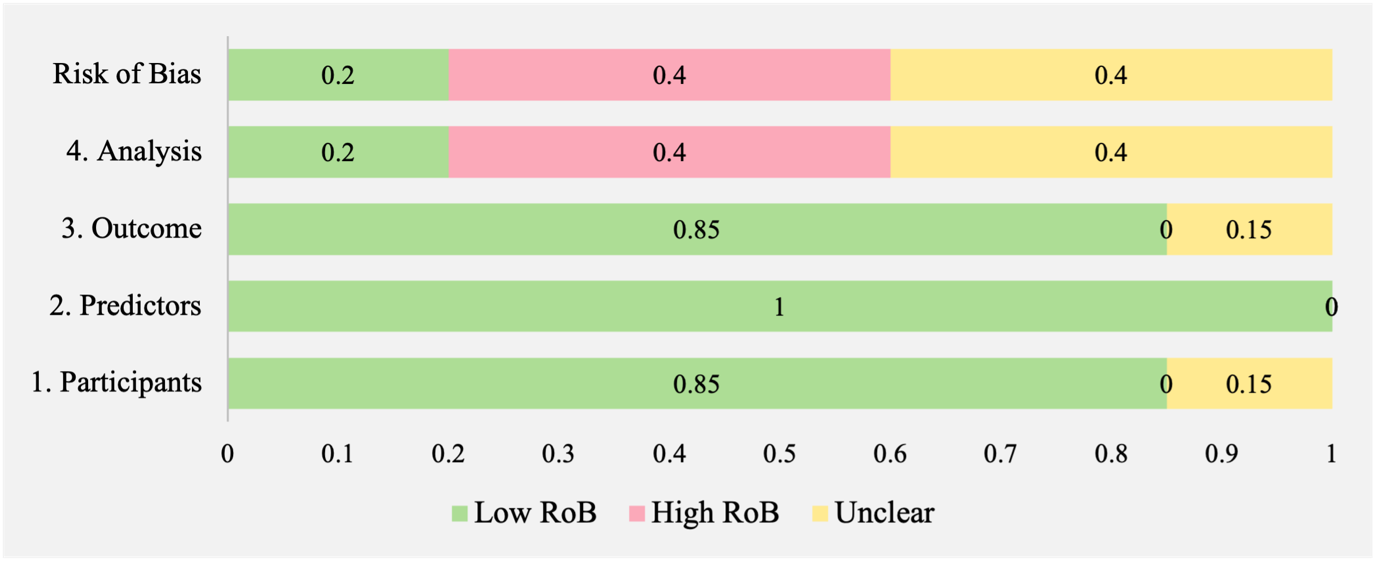

Abbreviations: RoB: Risk of Bias

**Supplementary Figure 3.** Summary of applicability assessment according to PROBAST (Prediction model Risk Of Bias ASsessment Tool)

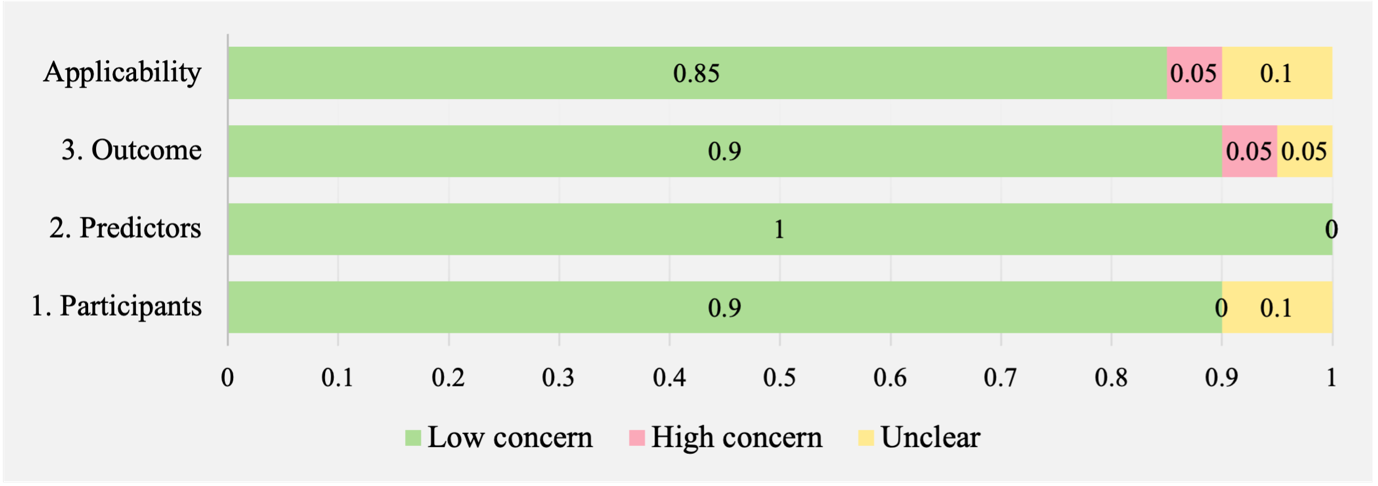

**Supplementary Tables**

**Supplementary Table 1.** PRISMA 2020 and PRISMA-S checklist^1^

| **Section and Topic** | **Item #** | **Checklist item** | **Location where item is reported** |
| --- | --- | --- | --- |
| **TITLE** | | |  |
| Title | 1 | Identify the report as a systematic review. | Title |
| **ABSTRACT** | | |  |
| Abstract | 2 | See the PRISMA 2020 for Abstracts checklist. | Not reported. |
| **INTRODUCTION** | | |  |
| Rationale | 3 | Describe the rationale for the review in the context of existing knowledge. | Introduction |
| Objectives | 4 | Provide an explicit statement of the objective(s) or question(s) the review addresses. | Introduction |
| **METHODS** | | |  |
| Eligibility criteria | 5 | Specify the inclusion and exclusion criteria for the review and how studies were grouped for the syntheses. | Methods – Study selection |
| Information sources | 6 | Specify all databases, registers, websites, organisations, reference lists and other sources searched or consulted to identify studies. Specify the date when each source was last searched or consulted. | Methods – Search Strategy |
| Search strategy | 7 | Present the full search strategies for all databases, registers and websites, including any filters and limits used. | Methods – Search Strategy and Supplementary Table 1 |
| Selection process | 8 | Specify the methods used to decide whether a study met the inclusion criteria of the review, including how many reviewers screened each record and each report retrieved, whether they worked independently, and if applicable, details of automation tools used in the process. | Methods – study selection criteria and screening process |
| Data collection process | 9 | Specify the methods used to collect data from reports, including how many reviewers collected data from each report, whether they worked independently, any processes for obtaining or confirming data from study investigators, and if applicable, details of automation tools used in the process. | Methods – data synthesis and analysis |
| Data items | 10a | List and define all outcomes for which data were sought. Specify whether all results that were compatible with each outcome domain in each study were sought (e.g. for all measures, time points, analyses), and if not, the methods used to decide which results to collect. | Methods – study selection criteria |
|  | 10b | List and define all other variables for which data were sought (e.g. participant and intervention characteristics, funding sources). Describe any assumptions made about any missing or unclear information. | Supplementary table 3 |
| Study risk of bias assessment | 11 | Specify the methods used to assess risk of bias in the included studies, including details of the tool(s) used, how many reviewers assessed each study and whether they worked independently, and if applicable, details of automation tools used in the process. | Methods – data synthesis and analysis |
| Effect measures | 12 | Specify for each outcome the effect measure(s) (e.g. risk ratio, mean difference) used in the synthesis or presentation of results. | Not reported. |
| Synthesis methods | 13a | Describe the processes used to decide which studies were eligible for each synthesis (e.g. tabulating the study intervention characteristics and comparing against the planned groups for each synthesis (item #5)). | Not reported. |
|  | 13b | Describe any methods required to prepare the data for presentation or synthesis, such as handling of missing summary statistics, or data conversions. | Not reported. |
|  | 13c | Describe any methods used to tabulate or visually display results of individual studies and syntheses. | Not reported. |
|  | 13d | Describe any methods used to synthesize results and provide a rationale for the choice(s). If meta-analysis was performed, describe the model(s), method(s) to identify the presence and extent of statistical heterogeneity, and software package(s) used. | Not reported. |
|  | 13e | Describe any methods used to explore possible causes of heterogeneity among study results (e.g. subgroup analysis, meta-regression). | Not reported. |
|  | 13f | Describe any sensitivity analyses conducted to assess robustness of the synthesized results. | Not reported. |
| Reporting bias assessment | 14 | Describe any methods used to assess risk of bias due to missing results in a synthesis (arising from reporting biases). | Methods – data synthesis and analysis |
| Certainty assessment | 15 | Describe any methods used to assess certainty (or confidence) in the body of eviNdence for an outcome. | Not reported. |
| **RESULTS** | | |  |
| Study selection | 16a | Describe the results of the search and selection process, from the number of records identified in the search to the number of studies included in the review, ideally using a flow diagram. | Results and Figure 1. |
|  | 16b | Cite studies that might appear to meet the inclusion criteria, but which were excluded, and explain why they were excluded. | Not applicable. |
| Study characteristics | 17 | Cite each included study and present its characteristics. | Table 1 |
| Risk of bias in studies | 18 | Present assessments of risk of bias for each included study. | Results – Quality and bias assessment, Supplementary table xx and Supplementary figure xx. |
| Results of individual studies | 19 | For all outcomes, present, for each study: (a) summary statistics for each group (where appropriate) and (b) an effect estimate and its precision (e.g. confidence/credible interval), ideally using structured tables or plots. | Supplementary table 4 and 5. |
| Results of syntheses | 20a | For each synthesis, briefly summarise the characteristics and risk of bias among contributing studies. | Results – Quality and bias assessment. |
|  | 20b | Present results of all statistical syntheses conducted. If meta-analysis was done, present for each the summary estimate and its precision (e.g. confidence/credible interval) and measures of statistical heterogeneity. If comparing groups, describe the direction of the effect. | Results – reported model performance. |
|  | 20c | Present results of all investigations of possible causes of heterogeneity among study results. | Not reported |
|  | 20d | Present results of all sensitivity analyses conducted to assess the robustness of the synthesized results. | Supplementary table S3-S4 |
| Reporting biases | 21 | Present assessments of risk of bias due to missing results (arising from reporting biases) for each synthesis assessed. | Results – Quality and bias assessment. |
| Certainty of evidence | 22 | Present assessments of certainty (or confidence) in the body of evidence for each outcome assessed. | Not reported |
| **DISCUSSION** | | |  |
| Discussion | 23a | Provide a general interpretation of the results in the context of other evidence. | Discussion |
|  | 23b | Discuss any limitations of the evidence included in the review. | Discussion – limitations |
|  | 23c | Discuss any limitations of the review processes used. | Limitations |
|  | 23d | Discuss implications of the results for practice, policy, and future research. | Discussion and conclusion |
| **OTHER INFORMATION** | | |  |
| Registration and protocol | 24a | Provide registration information for the review, including register name and registration number, or state that the review was not registered. | Not reported. |
|  | 24b | Indicate where the review protocol can be accessed, or state that a protocol was not prepared. | Supplementary table S1 |
|  | 24c | Describe and explain any amendments to information provided at registration or in the protocol. | Not performed. |
| Support | 25 | Describe sources of financial or non-financial support for the review, and the role of the funders or sponsors in the review. | Not applicable. |
| Competing interests | 26 | Declare any competing interests of review authors. | Acknowledgement section |
| Availability of data, code and other materials | 27 | Report which of the following are publicly available and where they can be found: template data collection forms; data extracted from included studies; data used for all analyses; analytic code; any other materials used in the review. | Not reported. |

| **Section and Topic** | **Item #** | **Checklist item** | **Location where item is reported** |
| --- | --- | --- | --- |
| INFORMATION SOURCES AND METHODS | | |  |
| Database name | 1 | Name each individual database searched, stating the platform for each. | Methods |
| Multi-database searching | 2 | If databases were searched simultaneously on a single platform, state the name of the platform, listing all of the databases searched. | Methods |
| Study registries | 3 | List any study registries searched. | Methods |
| Online resources and browsing | 4 | Describe any online or print source purposefully searched or browsed (e.g., tables of contents, print conference proceedings, web sites), and how this was done. | Not mentioned |
| Citation searching | 5 | Indicate whether cited references or citing references were examined, and describe any methods used for locating cited/citing references (e.g., browsing reference lists, using a citation index, setting up email alerts for references citing included studies). | Not done. |
| Contacts | 6 | Indicate whether additional studies or data were sought by contacting authors, experts, manufacturers, or others. | Methods |
| Other methods | 7 | Describe any additional information sources or search methods used. | Not done. |
| **SEARCH STRATEGIES** | | |  |
| Full search strategies | 8 | Include the search strategies for each database and information source, copied and pasted exactly as run. | Supplementary table 1 |
| Limits and restrictions | 9 | Specify that no limits were used, or describe any limits or restrictions applied to a search (e.g., date or time period, language, study design) and provide justification for their use. | Methods – search strategy |
| Search filters | 10 | Indicate whether published search filters were used (as originally designed or modified), and if so, cite the filter(s) used. | Methods – search strategy |
| Prior work | 11 | Indicate when search strategies from other literature reviews were adapted or reused for a substantive part or all of the search, citing the Not previous review(s). | Not done. |
| Updates | 12 | Report the methods used to update the search(es) (e.g., rerunning searches, email alerts). | Not done. |
| Dates of searches | 13 | For each search strategy, provide the date when the last search occurred. | Results |
| **PEER REVIEW** | | |  |
| Peer review | 14 | Describe any search peer review process. | Not done. |
| **MANAGING RECORDS** | | |  |
| Total Records | 15 | Document the total number of records identified from each database and other information sources. | Figure 1 |
| Deduplication | 16 | Describe the processes and any software used to deduplicate records from multiple database searches and other information sources. | Methods |

**Supplementary Table 2**. Search strategies used for Ovid MEDLINE (R)

| **n.** | **Search** |
| --- | --- |
|  | **Ovid MEDLINE(R)** |
| 1 | exp Electrocardiography/ |
| 2 | (electrocardiogra* or "ECG" or EKG or SAECG or "12-Lead ECG" or "twelve lead ECG").ti,ab,kf. |
| 3 | 1 or 2 |
| 4 | exp Machine Learning/ or exp Neural Networks, Computer/ or exp Pattern Recognition, Automated/ |
| 5 | ((machine or artificial* or deep or automat* or reinforce*) adj3 (model* or intelligence or reasoning* or rational* or learn* or detect* or algorithm*)).ti,ab,kf. |
| 6 | ("deep architec*" or "convolution* neural network*" or "comput* vision" or ResNet or DenseNEt or "convolution network" or ANN or "recurrent neural network" or "Long short-term memory" or LSTM).ti,ab,kf. |
| 7 | ((auto* or auto encod* or autoencod*) adj3 (quantif* or assess* or identificat* or detect* or classif*)).ti,ab,kf. |
| 8 | ((supervis* or automat* or algorit* or ensemble or transfer*) adj3 (classif* or learning)).ti,ab,kf. |
| 9 | ((neural or "generative adversarial") adj3 (network* or learn*)).ti,ab,kf. |
| 10 | 4 or 5 or 6 or 7 or 8 or 9 |
| 11 | 3 and 10 |
| 12 | ((exp animals/ or exp veterinary medicine/ or animal*.jw.) not exp humans/) or (experiment* model* or in vitro or animal* or monkey* or sheep or ?ovine or lamb* or goat* or pig* or swine or porcine or pup* or dog* or canine or bitch* or beagle* or feline or rodent* or rabbit* or rat or rats or mouse or murine or mice or horse or fish or bird or primate).ti,kf. |
| 13 | exp Case Reports/ or exp "Review"/ or exp Editorial/ or exp Letter/ or exp Comment/ or exp News/ |
| 14 | 12 or 13 |
| 15 | limit 11 to yr="2010 -Current" |
| 16 | 15 not 14 |

The search was conducted on October 5, 2023.

**Supplementary Table 3.** Extracted variables of the included articles

| **Category** | **Description** |
| --- | --- |
| Study design | Retrospective or prospective cohort. |
| Training data category | Clinical data, public data, or trial data. |
| Training data source | Description of database name and/or hospital name. |
| Training data period | Period in which included ECG-data was acquired. |
| Outcome definition | Definition used in study to determine the outcome left ventricular systolic dysfunction, including both modality and tresholding used. |
| Number of included samples | Number of included samples to train and/or (externally) validate developed models. This is determined both in number of patients and ECGs. |
| Subject inclusion | Defined inclusion criteria for eligibility in derivation and/or validation cohort. |
| Subject exclusion criteria | Defined exclusion criteria for eligibility in derivation and/or validation cohort. |
| Subject baseline characteristics | Reported characteristics on included subjects, including age, biological sex and ethnicity, |
| ECG data characteristics | Reported characteristics on included ECGs for derivation and/or external validation purposes, including acquisition device, sampling frequency, applied filtering strategy |
| Model input data | Data type used for the derivation of AI-ECG models, consisting of either signal-based or image-based input type. |
| Model architecture | Model architecture utilized to train described models. |
| Internal/External validation | Assessmsent of model performance in internal and/or external validation cohort. |
| Performance metrics | Reported performance metrics, including AUC, sensitivity, specificity, accuracy, PPV, and NPV reported in both internal and/or external validation cohorts. |
| Model explainability | Provided information on model explainability and feature importance techniques in the derivation study. |
| Bias assessment | Stratification for subgroups based on age, sex, or other demographic factors |
| Data availability | Reported data availability for study replication or use in future research, including: No data available, Data available upon request, Data available in published repository. |
| Code/Model availablity | Reported code and/or model availability for study replication and/or external model validation, including: No code available, Code available upon request, Model derivation code available, Full model and weights available |

Abbreviations: AUC, Area Under the receiver operating characteristics Curve; ECG, Electrocardiogram; NPV, Negative Predictive Value; PPV, Positive Predictive Value.

**Supplementary Table 4:** Characteristics of the derivation and internal validation cohorts in all included articles

| **First author** | **Year** | **Country** | **Outcome** | **No. ECGs** | **n outcome (%)** | **Age mean**  **± SD or**  **Median (iqr)** | **Female** | **External validation** |
| --- | --- | --- | --- | --- | --- | --- | --- | --- |
| Bergquist et al.^2^ | 2023 | USA | LVEF <40% | 24,868 | NR | NR | 45% | No |
| Khunte et al.^3^ | 2023 | USA | LVEF < 40% | 385,601 | 56,894 (14.8) | 68 (56-78) | 44% | No |
| Vaid et al.^4^ | 2023 | USA | LVEF < 40% | 511,491 | 94,114 (18.4) | NR | NR | Yes |
| Honarvar, Hossein^5^ | 2022 | USA | LVEF <35% | 92,446 | 7026(7.6) | 63.4 ± 15.1 | 45% | No |
| Huang, Yu-Chang^6^ | 2023 | Taiwan | LVEF < 40% | 380,675 | 8,216 (4.3) | 63.7 ± 16.3 | 46% | Yes |
| Chen, Hung-Yi^7^ | 2022 | Taiwan | LVEF < 35% | 88,597 | 5074 (5.7) | 66.5 ± 16.8 | 45% | Yes |
| Golany, Tomer^8^ | 2022 | Israel | LVEF < 50% | 13,820 | 6,910 (50) | 69.9 (59.7-80.2] | 39% | No |
| Yagi, Ryuichiro^9^ | 2022 | USA, Japan | LVEF <40% | 75,033 | NR | NR | NR | Yes |
| Lee et al.^10^ | 2022 | Taiwan | LVEF <40% | 127,912 | NR | 64.5 ± 17.2 | 49% | Yes |
| Sangha et al.^11^ | 2023 | USA | LVEF <40% | 385,601 | 56,895 (14.8) | 68 (56-78) | 51% | Yes |
| Chen et al.^12^ | 2022 | Taiwan | LVEF <35% | 127,440 | NR | 64.4 ± 17.2 | 49% | Yes |
| Kwon et al.^13^ | 2022 | Korea | LVEF <40% | 88,900 | 2,519 (6.5) | 59.3 ± 15.2 | 51% | Yes |
| Surendra et al.^14^ | 2023 | Germany | Asymptomatic LVSD, HFrEF (<35%), HFmrEF (<40%), HFpEF(>50%) | 5,299 | 318 (6.0) | 61.0 (54.0-68.0) | 49% | No |
| Vaid et al.^15^ | 2022 | USA | USA | 761,510 | 70,211 (9.2) | 65.6 (65.9-66.0) | 45% | Yes |
| Katsushika et al.^16^ | 2021 | Japan | LVEF <40% | 37,103 | 3505 (9.5) | 63.4 ± 16.9 | 43% | No |
| Cho et al.^17^ | 2021 | Korea | LVEF <40% | 42,841 | 2870 (6.7) | 60.8 ± 15.0 | 49% | Yes |
| Sun et al.^18^ | 2021 | China | LVEF <50% | 26,792 | 1,262 (4.7) | 56.6 ± 17.0 | 50% | No |
| Attia et al.^19^ | 2019 | USA | LVEF <35% | 97,829 | 7630 (7.8) | 54.1 ± 9.7 | 56% | No |
| van de Leur et al.^20^ | 2022 | The Netherlands | LVEF <40% | 39,603 | NR (13% of validation) | NR | NR | Yes |
| Vaid et al.^21^ | 2022 | USA | LVEF <40% | 723,701 | 135,295 (19) | 66 (66-66] | 42% | No |

Abbreviations: ECG: Electrocardiogram, NR: Not reported, SD: Standard deviation, iqr: inter quartile range.

**Supplementary Table 5:** Characteristics external validation cohorts in all included articles

| **First author** | **Year** | **Country** | **Outcome** | **Evaluated model** | **No. ECGs** | **No. Outcome (%)** | **Age mean**  **± SD or**  **Median (iqr)** | **Female (%)** |
| --- | --- | --- | --- | --- | --- | --- | --- | --- |
| Vaid et al.^4^ | 2023 | USA | LVEF < 40% | HeartBEiT, Vaid et al. | 1,480 | 394 (27) | NR | NR |
| Huang et al.^6^ | 2023 | Taiwan | LVEF < 40% | ResNet, Huang et al. | 91,425 | 2,812 (3) | 62 ± 17 | 45,245 (50) |
| Chen et al.^7^ | 2022 | Taiwan | LVEF <35% | ECG12Net <35%, Chen et al. | 20,629 | 742 (4) | 66 ± 17 | 10,259 (50) |
| Yagi et al.^9^ | 2022 | USA | LVEF <40% | CNN (BWH), Yaghi et al. | 79,663 | NR | NR | NR |
| Yagi et al.^9^ | 2022 | USA | LVEF <40% | CNN (BWH), Yaghi et al. | 36,314 | NR | NR | NR |
| Yagi et al.^9^ | 2022 | Japan | LVEF <40% | CNN (BWH), Yaghi et al. | 30,836 | NR | NR | NR |
| Yagi et al.^9^ | 2022 | USA | LVEF <40% | CNN (MGH), Yaghi et al. | 75,033 | NR | NR | NR |
| Yagi et al.^9^ | 2022 | Taiwan | LVEF <40% | ECG12NET (ALL SD), Lee et al. | 11,771 | NR | 66 ± 18 | 5,920 (501) |
| Sangha et al.^11^ | 2023 | USA | LVEF <40% | EfficientNet B3, Sangha et al. | 879 | 99 (11) | NR | NR |
| Sangha et al.^11^ | 2023 | USA | LVEF <40% | EfficientNet B3, Sangha et al. | 147 | 27 (18) | NR | NR |
| Sangha et al.^11^ | 2023 | USA | LVEF <40% | EfficientNet B3, Sangha et al. | 100 | 43 (43) | NR | NR |
| Sangha et al.^11^ | 2023 | USA | LVEF <40% | EfficientNet B3, Sangha et al. | 50 | 11 (22) | NR | NR |
| Sangha et al.^11^ | 2023 | USA | LVEF <40% | EfficientNet B3, Sangha et al. | 50 | 11 (20) | NR | NR |
| Sangha et al.^11^ | 2023 | Brazil | LVEF <40% | EfficientNet B3, Sangha et al. | 2,577 | 30 (1) | NR | NR |
| Chen et al.^12^ | 2022 | Taiwan | LVEF <35% | ECG12Net <35%, Chen et al. | 11,644 | NR | 66 ± 17 | 5,880 (51) |
| Kwon et al.^13^ | 2022 | Korea | LVEF <40% | ECGT2T <40%, Kwon et al. | 755 | 39 (5) | 56 ± 15 | 374 (50) |
| Vaid et al.^15^ | 2022 | USA | LVEF <35% | Efficientnet <35%, Vaid et al. | 1,439 | 332 (23) | 62.3 (NR) | 492 (34) |
| Cho et al.^17^ | 2021 | Korea | LVEF <40% | 12 lead CNN, Cho et al. | 4,362 |  |  |  |
| Choi et al.^22^ | 2022 | Korea | LVEF <40% | 12 lead CNN, Cho et al. | 1,291 | 547 (42) | 68 ± 14 | 568 (44) |
| Attia et al.^23^ | 2022 | USA | LVEF < 40% | CNN, Attia et al. | 421 | 16 (4) | 61 ± 18 | 195 (46) |
| Harmon et al.^24^ | 2022 | USA | LVEF < 50% | CNN, Attia et al. | 54 | 2 (4) | 55 (31;81) | 36 (67) |
| Harmon et al.^24^ | 2022 | USA | LVEF <40% | CNN, Attia et al. | 44,986 | 4,318 (10) | 64 ± 18 | 19,343 (43) |
| Attia et al.^25^ | 2022 | USA | LVEF < 35% | CNN, Attia et al. | 100 | 8 (8) | 61 ± 14 | 39 (39) |
| Attia et al.^26^ | 2021 | USA | LVEF <35% | CNN, Attia et al. | 4,277 | 26 (1) | 54 ± 10 | 2,395 (56) |
| Attia et al.^27^ | 2019 | USA | LVEF < 35% | CNN, Attia et al. | 3,874 | 271 (7) | 66 ± 15 | 2,196 (56) |
| Klein et al.^28^ | 2022 | USA | LVEF <35% | CNN, Attia et al. | 89 | NR | 62±13 | 43 (48) |
| Bachtiger et al.^29^ | 2021 | UK | LVEF < 40% | CNN, Attia et al. | 1,050 | 105 (10) | 62 ± 17 | 515 (49.0) |
| Kashou et al.^30^ | 2021 | USA | LVEF <35% | CNN, Attia et al. | 2,041 | 41 (2) | 36 ± 11 | 1,058 (52) |
| Brito et al.^31^ | 2021 | Brazil | LVEF <40% | CNN, Attia et al. | 1,304 | NR | 60 (51;69) | 872 (67) |
| Kashou et al.^32^ | 2021 | USA | LVEF 35% | CNN, Attia et al. | 528 | 310 (59) | 74 ± 12 | 193 (38) |
| Kashou et al.^32^ | 2021 | USA | LVEF 35% | CNN, Attia et al. | 3,252 | 2,244 (96) | 67 ± 15 | 1,120 (37) |
| Jentzer et al.^33^ | 2021 | USA | LVEF <35% | CNN, Attia et al. | 8,242 | 2,802 (34) | 68 ± 15 | 3,094 (37) |
| Adedinsewo et al.^34^ | 2020 | USA | LVEF <35% | CNN, Attia et al. | 1,606 | 164 (10) | 68 (57;78) | 759 (47.3) |
| Noseworthy et al.^35^ | 2020 | USA | LVEF <35% | CNN, Attia et al. | 97,829 | NR | 55 (NR) | 46.7 |
| van de Leur et al.^20^ | 2022 | UK | LVEF <40% | VAE + XGBoost, van de Leur et al. | 4855 | 28 (0.25%) | NR | NR |

Abbreviations: ECG, Electrocardiogram; NR, Not reported; SD, Standard deviation; iqr: inter quartile range.

**Supplementary Table 6:** Characteristics and internal performance of the models described in the included articles

| **First author** | **Year** | **Train/test/ Validation split** | **ECG type** | | **Model architecture** | | **AUROC [95%CI]** | **Reported threshold** | **Other reported performance metrics** | **Bias assessment** |
| --- | --- | --- | --- | --- | --- | --- | --- | --- | --- | --- |
| Bergquist, Jake A | 2023 | 90/10 | | 12 lead | | ResNet 18 | 0.92 [0.92;0.92] | Maximum F1 | Sensitivity, specificity, F1 | Age, Sex, Race, Comorbidities |
| Bergquist, Jake A | 2023 | 90/10 | | 12 lead | | ResNet 50 | 0.91 [0.91;0.92] | Maximum F1 | Sensitivity, specificity, F1 | Age, Sex, Race, Comorbidities |
| Bergquist, Jake A | 2023 | 90/10 | | 12 lead | | Alexnet | 0.92 [090;0.91] | Maximum F1 | Sensitivity, specificity, F1 | Age, Sex, Race, Comorbidities |
| Bergquist, Jake A | 2023 | 90/10 | | 12 lead | | Densenet 121 | 0.91 [0.90;0.91] | Maximum F1 | Sensitivity, specificity, F1 | Age, Sex, Race, Comorbidities |
| Bergquist, Jake A | 2023 | 90/10 | | 12 lead | | Sqeezenet | 0.90 [0.89;0.91] | Maximum F1 | Sensitivity, specificity, F1 | Age, Sex, Race, Comorbidities |
| Bergquist, Jake A | 2023 | 90/10 | | 12 lead | | VGG11 | 0.90 [0.89;0.90] | Maximum F1 | Sensitivity, specificity, F1 | Age, Sex, Race, Comorbidities |
| Khunte, Akshay | 2023 | 85/5/10 | | 1 lead | | CNN noise adapted | 0.90 [0.89;0.90] | Sensitivity 90% | Sensitivity, Specificity, NPV, PPV | Age, Sex, Race, noise |
| Khunte, Akshay | 2023 | 85/5/10 | | 1 lead | | CNN standard | 0.90 [0.88;0.90] | Sensitivity 90% | Sensitivity, Specificity, NPV, PPV | Age, Sex, Race, noise |
| Vaid, Akhil | 2023 | 96/3.8/0.2 | | 12 lead | | HeartBEiT | 0.90 [0.90;0.90] | NR | NR | No |
| Vaid, Akhil | 2023 | 96/3.8/0.2 | | 12 lead | | Vit-B/16 | 0.86 [0.86;0.86] | NR | NR | No |
| Vaid, Akhil | 2023 | 96/3.8/0.2 | | 12 lead | | EfficientNet B4 | 0.90 [0.90;0,90] | NR | NR | No |
| Vaid, Akhil | 2023 | 96/3.8/0.2 | | 12 lead | | Resnet 152 | 0.90 [0.90;0.90] | NR | NR | No |
| Honarvar, Hossein | 2022 | 80/10/10 | | 12 lead | | normal waveform representation CNN | 0.92 [NR] | NR | NR | Age, Sex, Race, ECG characteristics |
| Honarvar, Hossein | 2022 | 80/10/10 | | 12 lead | | Sub waveform representation CNN | 0.93 [NR] | NR | NR | Age, Sex, Race, ECG characteristics |
| Huang, Yu-Chang | 2023 | 35/15/50 | | 12 lead | | ResNet 18 signal | 0.95 [NR] | Youden's index | Accuracy, AUPRC, Sensitivity, Specificity, NPV, PPV | Age, Sex, Race, ECG characteristics |
| Huang, Yu-Chang | 2023 | 35/15/50 | | 12 lead | | Resnet 18 Image | 0.94 [NR]sc | Youden's index | Accuracy, AUPRC, Sensitivity, Specificity, NPV, PPV | Age, Sex, Race, ECG characteristics |
| Chen, Hung-Yi | 2022 | 64/36 | | 12 lead | | ECG12Net <35% | 0.93 [NR] | Youden's index | Sensitivity, Specificity | Age, Sex |
| Chen, Hung-Yi | 2022 | 64/36 | | 12 lead | | ECG12Net <50% | 0.88 [NR] | Youden's index | Sensitivity, Specificity | Age, Sex |
| Golany, Tomer | 2022 | 75/19/6 | | 12 lead | | ResNet <35% | 0.85 [NR] | Youden's index | Sensitivity, Specificity | No |
| Golany, Tomer | 2022 | 75/19/6 | | 12 lead | | ResNet <35% | 0.85 [NR] | Sensitivity 90% | Sensitivity, Specificity | No |
| Golany, Tomer | 2022 | 75/19/6 | | 12 lead | | ResNet <50% | 0.88 [NR] | Youden's index | Sensitivity, Specificity | No |
| Golany, Tomer | 2022 | 75/19/6 | | 12 lead | | ResNet <50% | 0.88 [NR] | Sensitivity 90% | Sensitivity, Specificity | No |
| Yagi, Ryuichiro | 2022 | 50/20/30 | | 12 lead | | CNN (BWH) | 0.91 [0.90;0.93] | NR | NR | Age, Sex, HR, ECG abnormalities |
| Yagi, Ryuichiro | 2022 | 50/20/30 | | 12 lead | | CNN ( MGH) | 0.87 [0.53;0.99] | NR | NR | Age, Sex, HR, ECG abnormalities |
| Yagi, Ryuichiro | 2022 | 50/20/30 | | 12 lead | | CNN (UCSF) | 0.90 [0.88;0,92] | NR | NR | Age, Sex, HR, ECG abnormalities |
| Yagi, Ryuichiro | 2022 | 50/20/30 | | 12 lead | | CNN (Keio) | 0.91 [0.89;0.94] | NR | NR | Age, Sex, HR, ECG abnormalities |
| Lee, Chun-Ho | 2022 | 80/10/10 | | 12 lead | | ECG12NET (ALL SD) | 0.95 [NR] | 0.212 | Sensitivity, Specificity, NPV, PPV | ECG characteristics |
| Lee, Chun-Ho | 2022 | 80/10/10 | | 12 lead | | ECG12NET (SD<12) | 0.96 [NR] | 0.212 | Sensitivity, Specificity, NPV, PPV | ECG characteristics |
| Lee, Chun-Ho | 2022 | 80/10/10 | | 12 lead | | ECG12NET(SD<10) | 0.9 [NR] | 0.212 | Sensitivity, Specificity, NPV, PPV | ECG characteristics |
| Sangha, Veer | 2023 | 85/5/10 | | 12 lead | | EfficientNet-B3 | 0.91 [0.90;0.92] | 90% sensitivity | Sensitivity, Specificity, NPV, PPV, F1 | Age, Sex, Race, AF, ECG characteristics, Paced rhythm |
| Chen, Hung-Yi | 2022 | 80/10/10 | | 12 lead | | ECG12Net | 0.96 [NR] | Youden's index | Sensitivity, Specificity, NPV, PPV | No |
| Chen, Hung-Yi | 2022 | 80/10/10 | | 12 lead | | ECG12Net | 0.88 [NR] | Youden's index | Sensitivity, Specificity, NPV, PPV | No |
| Kwon, Joon-Myoung | 2022 | 80/20 | | 2 lead | | ECGT2T <40% | 0.93 [0.93;0.94] | NR | NR | No |
| Kwon, Joon-Myoung | 2022 | 80/20 | | 2 lead | | ECGT2T <50% | 0.91 [0.90;0.94] | NR | NR | No |
| Surendra, Kishore | 2023 | 64/16/20 | | 12 lead | | CNN | 0.73 [0.71;0.74] | NR | Sensitivity, Specificity | No |
| Vaid, Akhil | 2022 | 85/15 | | 12 lead | | Efficientnet <35% | 0.95 [0.95;0.95] | Youden's index | Sensitivity, Specificity | No |
| Vaid, Akhil | 2022 | 85/15 | | 12 lead | | Efficientnet <40% | 0.94 [0.94;0.94] | Youden's index | Sensitivity, Specificity | No |
| Vaid, Akhil | 2022 | 85/15 | | 12 lead | | Efficientnet 40%-50% | 0.82 [0.81;0.83] | Youden's index | Sensitivity, Specificity | No |
| Vaid, Akhil | 2022 | 85/15 | | 12 lead | | Efficientnet >50% | 0.89 [0.89;0.89] | Youden's index | Sensitivity, Specificity | No |
| Katsushika, Susumu | 2021 | 64/16,5/19,5 | | 12 lead | | CNN A | 0.94 [0.93;0.95] | NR | Accuracy, AUPRC, Sensitivity, Specificity, NPV, PPV | No |
| Katsushika, Susumu | 2021 | 64/16,5/19,5 | | 12 lead | | CNN B | 0.94 [0.93;0.95] | NR | Accuracy, AUPRC, Sensitivity, Specificity, NPV, PPV | No |
| Katsushika, Susumu | 2021 | 64/16,5/19,5 | | 12 lead | | CNN C | 0.95 [0.94;0.95] | NR | Accuracy, AUPRC, Sensitivity, Specificity, NPV, PPV | No |
| Cho, Jinwoo | 2021 | 80/20 | | 12 lead | | CNN 12 lead | 0.91 [0.90;0.93] | 90% sensitivity | Accuracy, AUPRC, Sensitivity, Specificity | No |
| Cho, Jinwoo | 2021 | 80/20 | | 12 lead | | CNN 1 lead | 0.87 [0.86;0.89] | 90% sensitivity | Accuracy, AUPRC, Sensitivity, Specificity | No |
| Sun, Jin-Yu | 2021 | 90/10/10 | | 12 lead | | CNN | 0.709 [NR] | NR | Accuracy, AUPRC, Sensitivity, Specificity, NPV, PPV | No |
| Attia, Zachi I | 2019 | 10/40/50 | | 12 lead | | CNN | 0.932 [NR] | Youden's index | Accuracy, AUPRC, Sensitivity, Specificity, NPV, F1 | Age, Sex, Comorbidities |
| Attia, Zachi I | 2019 | 10/40/50 | | 12 lead | | CNN | 0.932 [NR] | 90% sensitivity | Accuracy, AUPRC, Sensitivity, Specificity, NPV, F1 | Age, Sex, Comorbidities |
| van de Leur, Rutger R | 2022 | 75/25 | | 12 lead | | VAE with XGBoost | 0.890 [0.89;0.91] | NR | NR | No |
| Vaid, Akhil | 2022 | 75/25 | | 12 lead | | EfficientNet B3 | 0.860 [0.83;0.88] | Youden's index | Sensitivity, Specificity | No |
| Vaid, Akhil | 2022 | 75/25 | | 12 lead | | EfficientNet B3 | 0.680 [0.63;0.73] | Youden's index | Sensitivity, Specificity | No |
| Vaid, Akhil | 2022 | 75/25 | | 12 lead | | EfficientNet B3 | 0.830 [0.8;0.85] | Youden's index | Sensitivity, Specificity | No |

Abbreviations: AUROC, area under the receiver operating characteristic curve; AUPRC, area under the precision-recall curve; CNN, convolutional neural network; VAE, variational autoencoder; HR, Hazart ratio; NPV, negative predictive value; PPV, positive predictive value; F1, F1 score; ResNet, residual network; VGG: visual geometry group network; SD, standard deviation; ECG, electrocardiogram.

**Supplementary Table 7.** External performance of the models described in the included articles

| **First author** | **Year** | **Evaluated model** | **AUROC [95%CI]** | **Other reported performance metrics** | **Bias assessment** |
| --- | --- | --- | --- | --- | --- |
| Vaid, Akhil | 2023 | HeartBEiT, Vaid et al. | 0.93 [0.93;0.93] | AUPRC | NR |
| Vaid, Akhil | 2023 | Vit-B/16, Vaid et al. | 0.89 [0.89;0.89] | AUPRC | NR |
| Vaid, Akhil | 2023 | EfficientNet B4, Vaid et al. | 0.92 [0.92;0.92] | AUPRC | NR |
| Vaid, Akhil | 2023 | Resnet 152, Vaid et al. | 0.92 [0.93;0.93] | AUPRC | NR |
| Huang, Yu-Chang | 2023 | ResNet, Huang et al. | 0.95 [NR] | Accuracy, Sensitivity, Specificity, NPV, PPV | Age, Sex, Race, ECG characteristics |
| Chen, Hung-Yi | 2022 | ECG12Net <35%, Chen et al. | 0.95 [NR] | Sensitivity, Specificity, | Age, Sex |
| Chen, Hung-Yi | 2022 | ECG12Net <50%, Chen et al. | 0.88 [NR] | Sensitivity, Specificity, | Age, Sex |
| Yagi, Ryuichiro | 2022 | CNN (BWH), Yaghi et al. | 0.9 [0.88;0.93] | NR | Age, Sex, Heart Rate, ECG abnormalities |
| Yagi, Ryuichiro | 2022 | CNN (BWH), Yaghi et al. | 0.91 [0.84;0.93] | NR | NR |
| Yagi, Ryuichiro | 2022 | CNN (BWH), Yaghi et al. | 0.92 [0.9;0.94] | NR | NR |
| Yagi, Ryuichiro | 2022 | CNN (MGH), Yaghi et al. | 0.9 [0.88;0.91] | NR | NR |
| Yagi, Ryuichiro | 2022 | CNN (MGH), Yaghi et al. | 0.89 [0.87;0.91] | NR | NR |
| Yagi, Ryuichiro | 2022 | CNN (MGH), Yaghi et al. | 0.92 [0.9;0.94] | NR | NR |
| Yagi, Ryuichiro | 2022 | CNN (UCSF), Yaghi et al. | 0.89 [0.88;0.9] | NR | NR |
| Yagi, Ryuichiro | 2022 | CNN (UCSF), Yaghi et al. | 0.87 [0.85;0.89] | NR | NR |
| Yagi, Ryuichiro | 2022 | CNN (UCSF), Yaghi et al. | 0.92 [0.9;0.94] | NR | NR |
| Yagi, Ryuichiro | 2022 | CNN (Keio), Yaghi et al. | 0.85 [0.83;0.83] | NR | NR |
| Yagi, Ryuichiro | 2022 | CNN (Keio), Yaghi et al. | 0.88 [0.86;0.9] | NR | NR |
| Yagi, Ryuichiro | 2022 | CNN (Keio), Yaghi et al. | 0.86 [0.84;0.88] | NR | NR |
| Lee, Chun-Ho | 2022 | ECG12NET (ALL SD), Lee et al. | 0.94 [NR] | Sensitivity, Specificity, NPV, PPV | AF, SR, HR, Printerval, QRS duration, QTc, Pwave axis, QRS axis, T axis, Paced |
| Lee, Chun-Ho | 2022 | ECG12NET (SD<12) , Lee et al. | 0.95 [NR] | Sensitivity, Specificity, NPV, PPV | AF, SR, HR, Printerval, QRS duration, QTc, Pwave axis, QRS axis, T axis, Paced |
| Lee, Chun-Ho | 2022 | ECG12NET(SD<10) , Lee et al. | 0.97 [NR] | Sensitivity, Specificity, NPV, PPV | AF, SR, HR, Printerval, QRS duration, QTc, Pwave axis, QRS axis, T axis, Paced |
| Sangha, Veer | 2023 | EfficientNet B3, Sangha et al. | 0.9 [0.88;0.93] | AUPRC, Sensitivity, Specificity, NPV, PPV, F1 | Age, Sex, Race, ECG findings, AF, Paced |
| Sangha, Veer | 2023 | EfficientNet B3, Sangha et al. | 0.95 [0.91;0.98] | AUPRC, Sensitivity, Specificity, NPV, PPV, F1 | Age, Sex, Race, ECG findings, AF, Paced |
| Sangha, Veer | 2023 | EfficientNet B3, Sangha et al. | 0.9 [0.84;0.96] | AUPRC, Sensitivity, Specificity, NPV, PPV, F1 | Age, Sex, Race, ECG findings, AF, Paced |
| Sangha, Veer | 2023 | EfficientNet B3, Sangha et al. | 0.92 [0.79;1.0] | AUPRC, Sensitivity, Specificity, NPV, PPV, F1 | Age, Sex, Race, ECG findings, AF, Paced |
| Sangha, Veer | 2023 | EfficientNet B3, Sangha et al. | 0.9 [0.82;0.99] | AUPRC, Sensitivity, Specificity, NPV, PPV, F1 | Age, Sex, Race, ECG findings, AF, Paced |
| Sangha, Veer | 2023 | EfficientNet B3, Sangha et al. | 0.95 [0.92;0.98] | AUPRC, Sensitivity, Specificity, NPV, PPV, F1 | Age, Sex, Race, ECG findings, AF, Paced |
| Chen, Hung-Yi | 2022 | ECG12Net <35%, Chen et al. | 0.94 [NR] | Sensitivity, Specificity, NPV, PPV | NR |
| Chen, Hung-Yi | 2022 | ECG12Net <50%, Chen et al. | 0.88 [NR] | Sensitivity, Specificity, NPV, PPV | NR |
| Kwon, Joon-Myoung | 2022 | ECGT2T <40%, Kwon et al. | 0.93 [0.91;0.96] | Sensitivity, Specificity, NPV, PPV | NR |
| Kwon, Joon-Myoung | 2022 | ECGT2T <50%, Kwon et al. | 0.84 [0.8;0.88] | Sensitivity, Specificity, NPV, PPV | NR |
| Vaid, Akhil | 2022 | Efficientnet <35%, Vaid et al. | 0.95 [0.95;0.96] | AUPRC, Sensitivity, Specificity | NR |
| Vaid, Akhil | 2022 | Efficientnet <40%, Vaid et al. | 0.94 [0.94;0.95] | AUPRC, Sensitivity, Specificity | NR |
| Vaid, Akhil | 2022 | Efficientnet 40%-50%, Vaid et al. | 0.73 [0.72;0.74] | AUPRC, Sensitivity, Specificity | NR |
| Vaid, Akhil | 2022 | Efficientnet >50%, Vaid et al. | 0.87 [0.87;0.88] | AUPRC, Sensitivity, Specificity | NR |
| Cho, Jinwoo | 2021 | 12 lead CNN, Cho et al. | 0.96 [0.95;0.97] | Sensitivity, Specificity | NR |
| Cho, Jinwoo | 2021 | 1 lead CNN, Cho et al. | 0.93 [0.91;0.96] | Sensitivity, Specificity | NR |
| Choi, JungMin | 2022 | 12 lead CNN, Cho et al. | 0.84 [0.82;0.87] | Accuracy, Sensitivity, Specificity, NPV, PPV | Age, Sex, DM, denovo versus acute HF, ischemic, BNP, eGFR, HR, SR or AF, PRinterval, QRSduration, Q-wave, QRS axis |
| Attia, Zachi I | 2022 | CNN, Attia et al. | 0.88 [0.82;0.95] | Sensitivity, Specificity | AF |
| Attia, Zachi I | 2022 | CNN, Attia et al. | 0.88 [0.82;0.95] |  | AF |
| Harmon, David M | 2022 | CNN, Attia et al. | 0.89 [0.71;1.0] | Sensitivity, Specificity | NR |
| Harmon, David M | 2022 | CNN, Attia et al. | 0.9 [0.9;0.91] | Sensitivity, Specificity | Sex, Age, Race, LVEF modality, Clinical care setting, Site location |
| Attia, Zachi | 2022 | CNN, Attia et al. | 0.86 [0.69;1.0] | Sensitivity, Specificity, NPV, PPV | Common comorbidities: AF, dyspnea, HF, syncope, bradycardia, arrhytmia, other |
| Attia, Zachi | 2022 | CNN, Attia et al. | 0.85 [0.71;0.95] | Sensitivity, Specificity, NPV, PPV | Common comorbidities: AF, dyspnea, HF, syncope, bradycardia, arrhytmia, other |
| Attia, Zachi | 2022 | CNN, Attia et al. | 0.83 [0.71;0.95] | Sensitivity, Specificity, NPV, PPV | Common comorbidities: AF, dyspnea, HF, syncope, bradycardia, arrhytmia, other |
| Attia, Itzhak Zachi | 2021 | CNN, Attia et al. | 0.82 [0.75;0.9] | Accuracy, Sensitivity, Specificity, NPV, PPV, F1 | Age, sex |
| Attia, Zachi I | 2019 | CNN, Attia et al. | 0.92 [0.9;0.93] | Accuracy, Sensitivity, Specificity | Race |
| Klein, Christopher J | 2022 | CNN, Attia et al. | 0.74 [0.6;0.87] | Sensitivity, Specificity | NR |
| Bachtiger, Patrik | 2021 | CNN, Attia et al. | 0.85 [0.81;0.89] | Sensitivity, Specificity, NPV, PPV, F1 | Age, Sex, Race |
| Bachtiger, Patrik | 2021 | CNN, Attia et al. | 0.91 [0.88;0.95] | Sensitivity, Specificity, NPV, PPV, F1 | Age, Sex, Race |
| Kashou, Anthony H | 2021 | CNN, Attia et al. | 0.98 [NR] | Sensitivity, Specificity | Age, Sex, Race |
| Kashou, Anthony H | 2021 | CNN, Attia et al. | 0.97 [NR] | Sensitivity, Specificity | Age, Sex, Race |
| Kashou, Anthony H | 2021 | CNN, Attia et al. | 0.88 [NR] | Sensitivity, Specificity | Age, Sex, Race |
| Brito, Bruno Oliveira | 2021 | CNN, Attia et al. | 0.84 [0.79;0.88] | Accuracy, Sensitivity, Specificity, NPV, PPV | Age, Sex, Race, disease history, ECG abnormalities |
| Kashou, Anthony H | 2021 | CNN, Attia et al. | 0.79 [NR] | Accuracy, Sensitivity, Specificity, NPV, PPV | AF, SR |
| Kashou, Anthony H | 2021 | CNN, Attia et al. | 0.82 [NR] | Accuracy, Sensitivity, Specificity, NPV, PPV | AF, SR |
| Jentzer, Jacob C | 2021 | CNN, Attia et al. | 0.83 [0.82;0.85] | Accuracy, Sensitivity, Specificity, NPV, PPV | Age, Sex, EF subgroups, measurement day |
| Adedinsewo, Demilade | 2020 | CNN, Attia et al. | 0.88 [0.86;0.91] | Accuracy, Sensitivity, Specificity, NPV, PPV, F1 | Age, Sex |
| Adedinsewo, Demilade | 2020 | CNN, Attia et al. | 0.85 [0.83;0.88] | Accuracy, Sensitivity, Specificity, NPV, PPV, F1 | Age, Sex |
| Noseworthy, Peter A | 2020 | CNN, Attia et al. | 0.93 [NR] | NR | Race |
| van de Leur, Rutger R | 2022 | VAE + XGBoost, van de Leur et al. | 0.89 [0.84;0.95] | AUPRC | NR |

Abbreviations: AUROC, area under the receiver operating characteristic curve; AUPRC, area under the precision-recall curve; CNN, convolutional neural network; VAE, variational autoencoder; HR, Hazart ratio; NPV, negative predictive value; PPV, positive predictive value; F1, F1 score; ResNet, residual network; VGG: visual geometry group network; SD, standard deviation; ECG, electrocardiogram.

**Supplementary table 8.** Performance of the four included AI-ECG models in the full CMR cohort

| **Model** | **Number of patients** | **Number of outcomes** | **AUROC** **[95%CI]** | **Sensitivity**  **[95%CI]** | **Specificity**  **[95%CI]** | **PPV**  **[95%CI]** | **NPV**  **[95%CI]** | **Missing (%)** |
| --- | --- | --- | --- | --- | --- | --- | --- | --- |
| **AiTiALVSD** | 1203 | 212 | 0.938 [0.933;0.943] | 0.879 [0.858;0.901] | 0.855 [0.848;0.865] | 0.565 [0.551;0.582] | 0.971 [0.966;0.976] | 0.00 |
| Male | 769 | 162 | 0.938 [0.932;0.943] | 0.915 [0.894;0.935] | 0.839 [0.828;0.851] | 0.602 [0.584;0.620] | 0.974 [0.967;0.980] | 0.00 |
| Age <60 | 537 | 67 | 0.939 [0.929;0.949] | 0.849 [0.806;0.888] | 0.894 [0.881;0.907] | 0.533 [0.500;0.567] | 0.976 [0.970;0.983] | 0.00 |
| Age ≥60 | 662 | 144 | 0.937 [0.931;0.943] | 0.900 [0.875;0.924] | 0.822 [0.810;0.834] | 0.582 [0.566;0.600] | 0.967 [0.960;0.975] | 0.00 |
| QRS <100 ms | 636 | 53 | 0.951 [0.933;0.964] | 0.799 [0.736;0.860] | 0.935 [0.924;0.947] | 0.530 [0.475;0.583] | 0.981 [0.975;0.987] | 0.00 |
| QRS ≥100 ms | 566 | 159 | 0.912 [0.902;0.922] | 0.906 [0.883;0.925] | 0.741 [0.726;0.758] | 0.577 [0.558;0.596] | 0.953 [0.942;0.963] | 0.00 |
| Heart Rate >80 bpm | 310 | 78 | 0.923 [0.906;0.940] | 0.892 [0.859;0.927] | 0.794 [0.761;0.826] | 0.592 [0.544;0.638] | 0.957 [0.943;0.970] | 0.00 |
| Heart Rate ≤80 bpm | 892 | 134 | 0.943 [0.932;0.951] | 0.871 [0.838;0.900] | 0.874 [0.865;0.885] | 0.550 [0.524;0.577] | 0.975 [0.968;0.981] | 0.00 |
| Atrial Fibrillation | 54 | 21 | 0.881 [0.836;0.918] | 0.908 [0.833;0.957] | 0.654 [0.545;0.758] | 0.623 [0.517;0.733] | 0.919 [0.857;0.963] | 0.00 |
| No Atrial Fibrillation | 1148 | 191 | 0.939 [0.933;0.945] | 0.876 [0.853;0.896] | 0.863 [0.854;0.871] | 0.559 [0.541;0.578] | 0.972 [0.967;0.977] | 0.00 |
| Coronary Disease | 327 | 83 | 0.908 [0.898;0.918] | 0.920 [0.890;0.952] | 0.768 [0.743;0.788] | 0.574 [0.549;0.599] | 0.966 [0.953;0.979] | 0.00 |
| Other CMP | 146 | 30 | 0.885 [0.870;0.899] | 0.760 [0.684;0.833] | 0.803 [0.780;0.822] | 0.494 [0.455;0.533] | 0.930 [0.912;0.949] | 0.00 |
| DCM | 83 | 64 | 0.944 [0.934;0.954] | 0.923 [0.899;0.938] | 0.737 [0.700;0.750] | 0.922 [0.908;0.924] | 0.743 [0.690;0.789] | 0.00 |
| Myocarditis | 48 | 10 | 0.883 [0.831;0.938] | 0.735 [0.600;0.900] | 0.790 [0.718;0.846] | 0.477 [0.389;0.571] | 0.920 [0.879;0.968] | 0.00 |
| HCM | 63 | 7 | 0.961 [0.941;0.972] | 1.000 [1.000;1.000] | 0.754 [0.714;0.786] | 0.337 [0.304;0.368] | 1.000 [1.000;1.000] | 0.00 |
| Ischemia | 87 | 8 | 0.957 [0.939;0.975] | 0.915 [0.857;1.000] | 0.870 [0.850;0.887] | 0.408 [0.353;0.471] | 0.991 [0.985;1.000] | 0.00 |
| Valvular Disease | 42 | 2 | 0.971 [0.950;0.988] | 1.000 [1.000;1.000] | 0.912 [0.864;0.951] | 0.371 [0.267;0.500] | 1.000 [1.000;1.000] | 0.00 |
| Congenital Heart Disease | 44 | 1 | 0.838 [0.814;0.884] | 0.000 [0.000;0.000] | 0.852 [0.814;0.907] | 0.000 [0.000;0.000] | 0.973 [0.972;0.975] | 0.00 |
| **ECG Vision** | 1203 | 212 | 0.885 [0.875;0.895] | 0.815 [0.791;0.838] | 0.816 [0.806;0.825] | 0.487 [0.470;0.504] | 0.954 [0.948;0.959] | 0.00 |
| Male | 769 | 162 | 0.881 [0.868;0.892] | 0.836 [0.809;0.859] | 0.786 [0.772;0.799] | 0.510 [0.492;0.527] | 0.947 [0.939;0.954] | 0.00 |
| Age <60 | 537 | 67 | 0.881 [0.862;0.898] | 0.730 [0.687;0.778] | 0.854 [0.840;0.866] | 0.417 [0.389;0.444] | 0.957 [0.950;0.964] | 0.00 |
| Age ≥60 | 662 | 144 | 0.885 [0.874;0.897] | 0.861 [0.833;0.888] | 0.783 [0.767;0.796] | 0.523 [0.504;0.542] | 0.953 [0.945;0.962] | 0.00 |
| QRS <100 ms | 636 | 53 | 0.884 [0.860;0.906] | 0.692 [0.628;0.755] | 0.908 [0.898;0.920] | 0.407 [0.360;0.457] | 0.970 [0.964;0.976] | 0.00 |
| QRS ≥100 ms | 566 | 159 | 0.845 [0.829;0.860] | 0.856 [0.831;0.881] | 0.685 [0.667;0.704] | 0.514 [0.494;0.535] | 0.924 [0.912;0.937] | 0.00 |
| Heart Rate >80 bpm | 310 | 78 | 0.881 [0.856;0.902] | 0.870 [0.828;0.909] | 0.745 [0.712;0.779] | 0.534 [0.486;0.579] | 0.945 [0.927;0.961] | 0.00 |
| Heart Rate ≤80 bpm | 892 | 134 | 0.884 [0.868;0.899] | 0.783 [0.746;0.814] | 0.838 [0.827;0.850] | 0.460 [0.434;0.489] | 0.956 [0.949;0.963] | 0.00 |
| Atrial Fibrillation | 54 | 21 | 0.846 [0.771;0.916] | 0.822 [0.725;0.909] | 0.684 [0.571;0.794] | 0.621 [0.500;0.750] | 0.861 [0.796;0.926] | 0.00 |
| No Atrial Fibrillation | 1148 | 191 | 0.886 [0.876;0.896] | 0.814 [0.790;0.835] | 0.821 [0.811;0.831] | 0.475 [0.459;0.494] | 0.957 [0.951;0.962] | 0.00 |
| Coronary Disease | 327 | 83 | 0.867 [0.849;0.886] | 0.854 [0.810;0.892] | 0.756 [0.730;0.783] | 0.544 [0.512;0.575] | 0.939 [0.921;0.955] | 0.00 |
| Other CMP | 146 | 30 | 0.818 [0.789;0.841] | 0.727 [0.655;0.800] | 0.740 [0.720;0.761] | 0.414 [0.380;0.451] | 0.915 [0.895;0.935] | 0.00 |
| DCM | 83 | 64 | 0.858 [0.823;0.874] | 0.862 [0.828;0.891] | 0.733 [0.700;0.750] | 0.916 [0.902;0.919] | 0.615 [0.560;0.667] | 0.00 |
| Myocarditis | 48 | 10 | 0.825 [0.755;0.887] | 0.761 [0.700;0.800] | 0.734 [0.658;0.795] | 0.428 [0.356;0.500] | 0.922 [0.897;0.939] | 0.00 |
| HCM | 63 | 7 | 0.861 [0.769;0.898] | 0.838 [0.714;0.857] | 0.601 [0.571;0.643] | 0.208 [0.179;0.231] | 0.968 [0.943;0.973] | 0.00 |
| Ischemia | 87 | 8 | 0.870 [0.789;0.931] | 0.729 [0.500;0.875] | 0.843 [0.818;0.867] | 0.312 [0.235;0.389] | 0.969 [0.944;0.986] | 0.00 |
| Valvular Disease | 42 | 2 | 0.826 [0.793;0.863] | 0.500 [0.500;0.500] | 0.939 [0.900;0.976] | 0.308 [0.200;0.500] | 0.974 [0.973;0.976] | 0.00 |
| Congenital Heart Disease | 44 | 1 | 0.721 [0.698;0.744] | 0.000 [0.000;0.000] | 0.802 [0.791;0.814] | 0.000 [0.000;0.000] | 0.972 [0.971;0.972] | 0.00 |
| **CGMH** | 1203 | 212 | 0.901 [0.893;0.908] | 0.840 [0.816;0.867] | 0.808 [0.799;0.817] | 0.483 [0.470;0.497] | 0.959 [0.953;0.966] | 0.00 |
| Male | 769 | 162 | 0.899 [0.889;0.908] | 0.866 [0.840;0.895] | 0.794 [0.783;0.806] | 0.528 [0.511;0.543] | 0.957 [0.949;0.966] | 0.00 |
| Age <60 | 537 | 67 | 0.910 [0.898;0.923] | 0.763 [0.716;0.809] | 0.852 [0.840;0.864] | 0.424 [0.400;0.449] | 0.962 [0.954;0.969] | 0.00 |
| Age ≥60 | 662 | 144 | 0.889 [0.878;0.899] | 0.881 [0.853;0.910] | 0.768 [0.757;0.779] | 0.512 [0.498;0.527] | 0.959 [0.950;0.969] | 0.00 |
| QRS <100 ms | 636 | 53 | 0.925 [0.906;0.942] | 0.672 [0.604;0.745] | 0.905 [0.894;0.916] | 0.392 [0.344;0.438] | 0.968 [0.960;0.976] | 0.00 |
| QRS ≥100 ms | 566 | 159 | 0.848 [0.832;0.861] | 0.896 [0.872;0.918] | 0.669 [0.652;0.686] | 0.513 [0.495;0.531] | 0.943 [0.930;0.955] | 0.00 |
| Heart Rate >80 bpm | 310 | 78 | 0.900 [0.881;0.917] | 0.886 [0.849;0.920] | 0.764 [0.735;0.796] | 0.557 [0.514;0.603] | 0.952 [0.938;0.966] | 0.00 |
| Heart Rate ≤80 bpm | 892 | 134 | 0.899 [0.886;0.910] | 0.813 [0.777;0.850] | 0.822 [0.811;0.833] | 0.445 [0.423;0.470] | 0.961 [0.954;0.969] | 0.00 |
| Atrial Fibrillation | 54 | 21 | 0.864 [0.806;0.921] | 0.902 [0.824;0.958] | 0.594 [0.485;0.688] | 0.582 [0.484;0.676] | 0.907 [0.841;0.957] | 0.00 |
| No Atrial Fibrillation | 1148 | 191 | 0.901 [0.891;0.909] | 0.833 [0.806;0.861] | 0.816 [0.806;0.825] | 0.473 [0.456;0.491] | 0.961 [0.955;0.967] | 0.00 |
| Coronary Disease | 327 | 83 | 0.877 [0.860;0.896] | 0.849 [0.807;0.893] | 0.785 [0.762;0.807] | 0.573 [0.547;0.603] | 0.939 [0.923;0.956] | 0.00 |
| Other CMP | 146 | 30 | 0.825 [0.798;0.848] | 0.754 [0.690;0.828] | 0.690 [0.667;0.709] | 0.381 [0.345;0.414] | 0.917 [0.898;0.942] | 0.00 |
| DCM | 83 | 64 | 0.845 [0.824;0.862] | 0.873 [0.844;0.892] | 0.629 [0.600;0.650] | 0.887 [0.873;0.892] | 0.597 [0.545;0.632] | 0.00 |
| Myocarditis | 48 | 10 | 0.907 [0.855;0.954] | 0.897 [0.800;1.000] | 0.843 [0.789;0.897] | 0.600 [0.513;0.692] | 0.970 [0.939;1.000] | 0.00 |
| HCM | 63 | 7 | 0.938 [0.865;0.977] | 0.981 [0.857;1.000] | 0.596 [0.554;0.625] | 0.233 [0.207;0.250] | 0.996 [0.971;1.000] | 0.00 |
| Ischemia | 87 | 8 | 0.895 [0.855;0.936] | 0.896 [0.857;1.000] | 0.815 [0.785;0.838] | 0.322 [0.273;0.364] | 0.988 [0.984;1.000] | 0.00 |
| Valvular Disease | 42 | 2 | 0.872 [0.839;0.900] | 0.500 [0.500;0.500] | 0.839 [0.800;0.865] | 0.134 [0.111;0.155] | 0.971 [0.970;0.972] | 0.00 |
| Congenital Heart Disease | 44 | 1 | 0.846 [0.814;0.884] | 1.000 [1.000;1.000] | 0.749 [0.721;0.791] | 0.085 [0.077;0.100] | 1.000 [1.000;1.000] | 0.00 |
| **Utrecht** | 1130 | 193 | 0.832 [0.820;0.845] | 0.890 [0.868;0.913] | 0.558 [0.544;0.572] | 0.293 [0.282;0.304] | 0.961 [0.953;0.968] | 6.02 |
| Male | 720 | 146 | 0.842 [0.826;0.856] | 0.906 [0.882;0.927] | 0.554 [0.536;0.572] | 0.341 [0.326;0.356] | 0.959 [0.948;0.967] | 6.02 |
| Age <60 | 515 | 61 | 0.810 [0.787;0.832] | 0.852 [0.805;0.885] | 0.559 [0.542;0.580] | 0.207 [0.192;0.222] | 0.965 [0.955;0.973] | 6.02 |
| Age ≥60 | 612 | 130 | 0.842 [0.827;0.856] | 0.908 [0.883;0.932] | 0.557 [0.539;0.576] | 0.357 [0.342;0.372] | 0.957 [0.946;0.968] | 6.02 |
| QRS <100 ms | 604 | 49 | 0.832 [0.802;0.860] | 0.833 [0.769;0.886] | 0.650 [0.634;0.668] | 0.174 [0.155;0.194] | 0.978 [0.968;0.984] | 6.02 |
| QRS ≥100 ms | 526 | 144 | 0.786 [0.766;0.804] | 0.910 [0.886;0.934] | 0.424 [0.403;0.446] | 0.372 [0.355;0.389] | 0.926 [0.907;0.945] | 6.02 |
| Heart Rate >80 bpm | 286 | 70 | 0.821 [0.789;0.847] | 0.916 [0.872;0.948] | 0.454 [0.420;0.485] | 0.352 [0.318;0.387] | 0.943 [0.915;0.964] | 6.02 |
| Heart Rate ≤80 bpm | 843 | 122 | 0.831 [0.813;0.848] | 0.876 [0.843;0.905] | 0.589 [0.573;0.604] | 0.265 [0.248;0.284] | 0.966 [0.956;0.974] | 6.02 |
| Atrial Fibrillation | 46 | 19 | 0.858 [0.775;0.927] | 0.991 [0.941;1.000] | 0.320 [0.211;0.438] | 0.498 [0.414;0.584] | 0.982 [0.875;1.000] | 6.02 |
| No Atrial Fibrillation | 1084 | 174 | 0.828 [0.814;0.841] | 0.880 [0.854;0.904] | 0.565 [0.552;0.579] | 0.279 [0.267;0.291] | 0.961 [0.953;0.968] | 6.02 |
| Coronary Disease | 302 | 74 | 0.823 [0.799;0.849] | 0.915 [0.879;0.946] | 0.530 [0.496;0.566] | 0.387 [0.362;0.416] | 0.951 [0.931;0.969] | 6.02 |
| Other CMP | 137 | 26 | 0.799 [0.764;0.827] | 0.915 [0.846;0.963] | 0.499 [0.473;0.527] | 0.302 [0.270;0.325] | 0.962 [0.932;0.983] | 6.02 |
| DCM | 79 | 61 | 0.741 [0.710;0.771] | 0.863 [0.825;0.902] | 0.366 [0.350;0.368] | 0.813 [0.797;0.824] | 0.459 [0.400;0.538] | 6.02 |
| Myocarditis | 44 | 9 | 0.841 [0.764;0.916] | 0.945 [0.875;1.000] | 0.449 [0.362;0.543] | 0.290 [0.233;0.347] | 0.972 [0.931;1.000] | 6.02 |
| HCM | 58 | 6 | 0.743 [0.655;0.785] | 0.993 [0.833;1.000] | 0.380 [0.343;0.426] | 0.152 [0.128;0.167] | 0.998 [0.952;1.000] | 6.02 |
| Ischemia | 79 | 7 | 0.917 [0.848;0.965] | 0.941 [0.750;1.000] | 0.565 [0.521;0.610] | 0.183 [0.132;0.216] | 0.990 [0.956;1.000] | 6.02 |
| Valvular Disease | 41 | 2 | 0.780 [0.750;0.810] | 0.500 [0.500;0.500] | 0.694 [0.634;0.769] | 0.077 [0.062;0.100] | 0.965 [0.962;0.968] | 6.02 |
| Congenital Heart Disease | 40 | 1 | 0.862 [0.810;0.900] | 1.000 [1.000;1.000] | 0.517 [0.487;0.538] | 0.050 [0.048;0.053] | 1.000 [1.000;1.000] | 6.02 |

Abbreviations: AUROC: area under the receiver operating curve, CI: confidence interval, PPV: positive predictive value, NPV: negative predictive value, CGMH: Chang Gung Memorial Hospital.

**Supplementary table 9.** Performance of the four included AI-ECG models in the representative HF cohort

| **Model** | **AUROC** **[95%CI]** | **Sensitivity** **[95%CI]** | **Specificity** **[95%CI]** | **PPV** **[95%CI]** | **NPV** **[95%CI]** | **Missing (%)** |
| --- | --- | --- | --- | --- | --- | --- |
| **AiTiALVSD** | 0.960 [0.942;0.975] | 0.889 [0.833;0.940] | 0.920 [0.904;0.934] | 0.665 [0.624;0.702] | 0.979 [0.969;0.989] | 0.00 |
| Male | 0.958 [0.938;0.976] | 0.914 [0.855;0.969] | 0.907 [0.884;0.927] | 0.696 [0.641;0.747] | 0.979 [0.965;0.992] | 0.00 |
| Female | 0.957 [0.920;0.987] | 0.807 [0.667;0.946] | 0.938 [0.914;0.958] | 0.569 [0.453;0.691] | 0.979 [0.963;0.995] | 0.00 |
| Age <60 | 0.956 [0.926;0.983] | 0.838 [0.727;0.954] | 0.934 [0.914;0.954] | 0.602 [0.500;0.709] | 0.980 [0.963;0.995] | 0.00 |
| Age ≥60 | 0.962 [0.943;0.980] | 0.919 [0.863;0.981] | 0.907 [0.883;0.928] | 0.695 [0.627;0.754] | 0.980 [0.965;0.995] | 0.00 |
| QRS <100 ms | 0.971 [0.950;0.990] | 0.836 [0.700;0.955] | 0.966 [0.953;0.978] | 0.608 [0.476;0.714] | 0.990 [0.982;0.997] | 0.00 |
| QRS ≥100 ms | 0.938 [0.911;0.963] | 0.903 [0.842;0.956] | 0.840 [0.807;0.869] | 0.681 [0.624;0.736] | 0.959 [0.935;0.980] | 0.00 |
| **ECG Vision** | 0.914 [0.889;0.936] | 0.824 [0.762;0.881] | 0.877 [0.862;0.889] | 0.544 [0.512;0.577] | 0.965 [0.953;0.977] | 0.00 |
| Male | 0.906 [0.873;0.933] | 0.842 [0.769;0.913] | 0.840 [0.819;0.859] | 0.550 [0.500;0.595] | 0.958 [0.940;0.976] | 0.00 |
| Female | 0.917 [0.871;0.962] | 0.764 [0.619;0.905] | 0.929 [0.907;0.951] | 0.521 [0.406;0.640] | 0.975 [0.957;0.989] | 0.00 |
| Age <60 | 0.902 [0.852;0.949] | 0.738 [0.600;0.870] | 0.889 [0.868;0.909] | 0.442 [0.348;0.536] | 0.966 [0.947;0.985] | 0.00 |
| Age ≥60 | 0.918 [0.890;0.945] | 0.870 [0.804;0.933] | 0.865 [0.842;0.889] | 0.598 [0.540;0.655] | 0.967 [0.949;0.982] | 0.00 |
| QRS <100 ms | 0.903 [0.851;0.954] | 0.706 [0.542;0.850] | 0.929 [0.916;0.942] | 0.383 [0.271;0.485] | 0.981 [0.968;0.989] | 0.00 |
| QRS ≥100 ms | 0.886 [0.853;0.917] | 0.857 [0.790;0.924] | 0.787 [0.753;0.817] | 0.602 [0.553;0.649] | 0.936 [0.907;0.965] | 0.00 |
| **CGMH** | 0.936 [0.916;0.955] | 0.864 [0.810;0.923] | 0.880 [0.864;0.894] | 0.563 [0.529;0.595] | 0.973 [0.962;0.985] | 0.00 |
| Male | 0.932 [0.911;0.952] | 0.896 [0.837;0.953] | 0.850 [0.828;0.869] | 0.582 [0.535;0.626] | 0.972 [0.958;0.987] | 0.00 |
| Female | 0.936 [0.885;0.979] | 0.761 [0.628;0.895] | 0.922 [0.899;0.946] | 0.500 [0.386;0.613] | 0.974 [0.957;0.989] | 0.00 |
| Age <60 | 0.939 [0.906;0.969] | 0.794 [0.667;0.906] | 0.903 [0.880;0.923] | 0.494 [0.385;0.587] | 0.973 [0.956;0.990] | 0.00 |
| Age ≥60 | 0.935 [0.908;0.963] | 0.903 [0.843;0.965] | 0.859 [0.835;0.883] | 0.597 [0.539;0.651] | 0.975 [0.959;0.991] | 0.00 |
| QRS <100 ms | 0.951 [0.917;0.978] | 0.704 [0.536;0.854] | 0.943 [0.929;0.956] | 0.432 [0.306;0.536] | 0.981 [0.969;0.993] | 0.00 |
| QRS ≥100 ms | 0.898 [0.866;0.929] | 0.909 [0.851;0.968] | 0.773 [0.735;0.803] | 0.602 [0.548;0.648] | 0.957 [0.933;0.985] | 0.00 |
| **Utrecht** | 0.865 [0.835;0.892] | 0.889 [0.836;0.943] | 0.634 [0.618;0.649] | 0.289 [0.270;0.310] | 0.972 [0.959;0.986] | 6.02 |
| Male | 0.862 [0.826;0.895] | 0.903 [0.841;0.957] | 0.606 [0.582;0.629] | 0.332 [0.296;0.363] | 0.967 [0.946;0.985] | 6.02 |
| Female | 0.855 [0.794;0.916] | 0.846 [0.706;0.952] | 0.677 [0.654;0.703] | 0.202 [0.140;0.261] | 0.978 [0.957;0.992] | 6.02 |
| Age <60 | 0.838 [0.772;0.899] | 0.843 [0.727;0.958] | 0.631 [0.609;0.654] | 0.196 [0.140;0.244] | 0.974 [0.957;0.993] | 6.02 |
| Age ≥60 | 0.878 [0.844;0.913] | 0.910 [0.846;0.962] | 0.636 [0.611;0.661] | 0.358 [0.318;0.398] | 0.969 [0.948;0.987] | 6.02 |
| QRS <100 ms | 0.860 [0.797;0.919] | 0.858 [0.730;1.000] | 0.706 [0.687;0.722] | 0.146 [0.104;0.188] | 0.988 [0.978;1.000] | 6.02 |
| QRS ≥100 ms | 0.823 [0.781;0.865] | 0.899 [0.841;0.960] | 0.512 [0.482;0.542] | 0.393 [0.356;0.429] | 0.935 [0.901;0.976] | 6.02 |

Abbreviations: AUROC: area under the receiver operating curve, CI: confidence interval, PPV: positive predictive value, NPV: negative predictive value, CGMH: Chang Gung Memorial Hospital.

**Supplementary table 10.** Performance of the four included AI-ECG models in the cohort without missing predictions.

| **Model** | **Number of patients** | **Number of outcome** | **AUROC** **[95%CI]** | **Sensitivity**  **[95%CI]** | **Specificity**  **[95%CI]** | **PPV**  **[95%CI]** | **NPV**  **[95%CI]** | **Missing**  **(%)** |
| --- | --- | --- | --- | --- | --- | --- | --- | --- |
| **AiTiALVSD** | 1171 | 206 | 0.941 [0.937;0.947] | 0.886 [0.864;0.905] | 0.858 [0.850;0.867] | 0.571 [0.554;0.589] | 0.972 [0.967;0.977] | 0.00 |
| Male | 747 | 157 | 0.943 [0.938;0.949] | 0.924 [0.904;0.949] | 0.842 [0.832;0.855] | 0.608 [0.591;0.631] | 0.977 [0.970;0.984] | 0.00 |
| Age <60 | 529 | 65 | 0.939 [0.928;0.949] | 0.845 [0.800;0.885] | 0.893 [0.881;0.905] | 0.526 [0.495;0.557] | 0.976 [0.970;0.982] | 0.00 |
| Age ≥60 | 638 | 140 | 0.943 [0.937;0.948] | 0.911 [0.892;0.935] | 0.826 [0.814;0.839] | 0.595 [0.575;0.614] | 0.971 [0.965;0.978] | 0.00 |
| QRS <100 ms | 625 | 52 | 0.952 [0.933;0.965] | 0.795 [0.729;0.859] | 0.936 [0.925;0.947] | 0.532 [0.481;0.588] | 0.980 [0.974;0.987] | 0.00 |
| QRS ≥100 ms | 545 | 154 | 0.918 [0.908;0.929] | 0.917 [0.895;0.936] | 0.744 [0.729;0.758] | 0.583 [0.564;0.600] | 0.958 [0.947;0.967] | 0.00 |
| Heart Rate >80 bpm | 301 | 75 | 0.923 [0.907;0.940] | 0.889 [0.854;0.923] | 0.799 [0.770;0.828] | 0.595 [0.546;0.644] | 0.956 [0.942;0.969] | 0.00 |
| Heart Rate ≤80 bpm | 869 | 131 | 0.948 [0.937;0.956] | 0.884 [0.849;0.916] | 0.876 [0.864;0.888] | 0.558 [0.530;0.585] | 0.977 [0.970;0.983] | 0.00 |
| Atrial Fibrillation | 50 | 21 | 0.883 [0.836;0.919] | 0.917 [0.853;0.958] | 0.670 [0.564;0.783] | 0.667 [0.571;0.760] | 0.918 [0.857;0.958] | 0.00 |
| No Atrial Fibrillation | 1120 | 185 | 0.942 [0.937;0.949] | 0.882 [0.859;0.903] | 0.864 [0.855;0.873] | 0.561 [0.543;0.581] | 0.974 [0.969;0.978] | 0.00 |
| Coronary Disease | 320 | 82 | 0.915 [0.904;0.925] | 0.929 [0.896;0.952] | 0.769 [0.748;0.794] | 0.581 [0.553;0.609] | 0.969 [0.955;0.979] | 0.00 |
| Other CMP | 139 | 27 | 0.894 [0.877;0.908] | 0.774 [0.698;0.852] | 0.805 [0.781;0.825] | 0.484 [0.442;0.524] | 0.938 [0.918;0.958] | 0.00 |
| DCM | 83 | 64 | 0.944 [0.932;0.953] | 0.922 [0.892;0.938] | 0.736 [0.700;0.750] | 0.921 [0.908;0.924] | 0.739 [0.667;0.778] | 0.00 |
| Myocarditis | 46 | 9 | 0.875 [0.823;0.937] | 0.708 [0.556;0.889] | 0.795 [0.737;0.842] | 0.453 [0.357;0.538] | 0.919 [0.879;0.968] | 0.00 |
| HCM | 60 | 6 | 0.968 [0.951;0.975] | 1.000 [1.000;1.000] | 0.763 [0.722;0.796] | 0.320 [0.286;0.353] | 1.000 [1.000;1.000] | 0.00 |
| Ischemia | 81 | 8 | 0.961 [0.940;0.977] | 0.923 [0.857;1.000] | 0.886 [0.865;0.905] | 0.461 [0.400;0.533] | 0.991 [0.984;1.000] | 0.00 |
| Valvular Disease | 42 | 2 | 0.972 [0.950;0.988] | 1.000 [1.000;1.000] | 0.914 [0.864;0.951] | 0.377 [0.267;0.500] | 1.000 [1.000;1.000] | 0.00 |
| Congenital Heart Disease | 43 | 1 | 0.825 [0.810;0.857] | 0.000 [0.000;0.000] | 0.836 [0.810;0.881] | 0.000 [0.000;0.000] | 0.972 [0.971;0.974] | 0.00 |
| **ECG Vision** | 1171 | 206 | 0.886 [0.875;0.895] | 0.810 [0.785;0.834] | 0.818 [0.809;0.827] | 0.487 [0.472;0.503] | 0.953 [0.947;0.958] | 0.00 |
| Male | 747 | 157 | 0.883 [0.870;0.894] | 0.830 [0.801;0.857] | 0.788 [0.775;0.801] | 0.510 [0.493;0.529] | 0.946 [0.938;0.954] | 0.00 |
| Age <60 | 529 | 65 | 0.878 [0.857;0.896] | 0.724 [0.677;0.773] | 0.853 [0.840;0.865] | 0.409 [0.383;0.432] | 0.957 [0.949;0.964] | 0.00 |
| Age ≥60 | 638 | 140 | 0.888 [0.876;0.899] | 0.856 [0.827;0.885] | 0.787 [0.774;0.801] | 0.529 [0.511;0.547] | 0.951 [0.942;0.960] | 0.00 |
| QRS <100 ms | 625 | 52 | 0.884 [0.859;0.906] | 0.684 [0.620;0.741] | 0.913 [0.901;0.925] | 0.417 [0.371;0.464] | 0.969 [0.963;0.976] | 0.00 |
| QRS ≥100 ms | 545 | 154 | 0.845 [0.831;0.861] | 0.853 [0.825;0.877] | 0.681 [0.662;0.698] | 0.511 [0.492;0.531] | 0.922 [0.908;0.934] | 0.00 |
| Heart Rate >80 bpm | 301 | 75 | 0.882 [0.859;0.903] | 0.864 [0.821;0.901] | 0.753 [0.723;0.782] | 0.537 [0.491;0.581] | 0.944 [0.928;0.959] | 0.00 |
| Heart Rate ≤80 bpm | 869 | 131 | 0.884 [0.870;0.899] | 0.779 [0.742;0.814] | 0.838 [0.828;0.850] | 0.460 [0.435;0.487] | 0.955 [0.948;0.962] | 0.00 |
| Atrial Fibrillation | 50 | 21 | 0.844 [0.773;0.903] | 0.823 [0.722;0.909] | 0.639 [0.518;0.750] | 0.622 [0.516;0.729] | 0.834 [0.750;0.913] | 0.00 |
| No Atrial Fibrillation | 1120 | 185 | 0.886 [0.875;0.896] | 0.808 [0.784;0.834] | 0.824 [0.814;0.833] | 0.476 [0.459;0.494] | 0.956 [0.951;0.962] | 0.00 |
| Coronary Disease | 320 | 82 | 0.870 [0.853;0.887] | 0.854 [0.815;0.892] | 0.755 [0.730;0.780] | 0.545 [0.515;0.574] | 0.938 [0.922;0.953] | 0.00 |
| Other CMP | 139 | 27 | 0.808 [0.774;0.833] | 0.695 [0.615;0.778] | 0.738 [0.717;0.761] | 0.386 [0.344;0.422] | 0.911 [0.890;0.933] | 0.00 |
| DCM | 83 | 64 | 0.856 [0.818;0.871] | 0.861 [0.828;0.891] | 0.735 [0.700;0.750] | 0.916 [0.902;0.919] | 0.612 [0.560;0.667] | 0.00 |
| Myocarditis | 46 | 9 | 0.802 [0.720;0.870] | 0.739 [0.667;0.778] | 0.731 [0.653;0.789] | 0.400 [0.333;0.467] | 0.921 [0.893;0.938] | 0.00 |
| HCM | 60 | 6 | 0.870 [0.758;0.907] | 0.812 [0.667;0.833] | 0.601 [0.574;0.630] | 0.184 [0.148;0.200] | 0.966 [0.939;0.971] | 0.00 |
| Ischemia | 81 | 8 | 0.873 [0.796;0.927] | 0.733 [0.500;0.875] | 0.858 [0.836;0.880] | 0.353 [0.267;0.425] | 0.968 [0.942;0.985] | 0.00 |
| Valvular Disease | 42 | 2 | 0.822 [0.787;0.854] | 0.500 [0.500;0.500] | 0.940 [0.900;0.976] | 0.318 [0.200;0.500] | 0.974 [0.973;0.976] | 0.00 |
| Congenital Heart Disease | 43 | 1 | 0.723 [0.714;0.738] | 0.000 [0.000;0.000] | 0.818 [0.810;0.833] | 0.000 [0.000;0.000] | 0.972 [0.971;0.972] | 0.00 |
| **CGMH** | 1171 | 206 | 0.902 [0.894;0.909] | 0.834 [0.810;0.859] | 0.813 [0.805;0.821] | 0.487 [0.474;0.500] | 0.958 [0.952;0.964] | 0.00 |
| Male | 747 | 157 | 0.900 [0.891;0.909] | 0.861 [0.833;0.886] | 0.801 [0.790;0.812] | 0.534 [0.517;0.552] | 0.956 [0.948;0.964] | 0.00 |
| Age <60 | 529 | 65 | 0.909 [0.896;0.922] | 0.756 [0.708;0.800] | 0.854 [0.841;0.866] | 0.421 [0.397;0.444] | 0.961 [0.954;0.968] | 0.00 |
| Age ≥60 | 638 | 140 | 0.891 [0.881;0.901] | 0.876 [0.843;0.906] | 0.774 [0.763;0.785] | 0.521 [0.503;0.536] | 0.957 [0.946;0.967] | 0.00 |
| QRS <100 ms | 625 | 52 | 0.926 [0.907;0.942] | 0.665 [0.588;0.735] | 0.910 [0.899;0.919] | 0.402 [0.356;0.448] | 0.968 [0.961;0.974] | 0.00 |
| QRS ≥100 ms | 545 | 154 | 0.848 [0.836;0.861] | 0.891 [0.869;0.915] | 0.672 [0.654;0.687] | 0.515 [0.496;0.533] | 0.940 [0.928;0.953] | 0.00 |
| Heart Rate >80 bpm | 301 | 75 | 0.902 [0.883;0.920] | 0.882 [0.846;0.915] | 0.777 [0.749;0.806] | 0.567 [0.527;0.610] | 0.952 [0.937;0.967] | 0.00 |
| Heart Rate ≤80 bpm | 869 | 131 | 0.899 [0.888;0.911] | 0.806 [0.766;0.838] | 0.824 [0.814;0.834] | 0.448 [0.425;0.469] | 0.960 [0.952;0.967] | 0.00 |
| Atrial Fibrillation | 50 | 21 | 0.853 [0.794;0.910] | 0.904 [0.842;0.957] | 0.597 [0.492;0.691] | 0.617 [0.519;0.710] | 0.897 [0.833;0.952] | 0.00 |
| No Atrial Fibrillation | 1120 | 185 | 0.901 [0.894;0.909] | 0.826 [0.800;0.851] | 0.820 [0.811;0.828] | 0.475 [0.460;0.491] | 0.960 [0.954;0.965] | 0.00 |
| Coronary Disease | 320 | 82 | 0.883 [0.868;0.898] | 0.844 [0.805;0.884] | 0.794 [0.774;0.813] | 0.584 [0.556;0.609] | 0.937 [0.921;0.952] | 0.00 |
| Other CMP | 139 | 27 | 0.819 [0.791;0.842] | 0.726 [0.654;0.804] | 0.698 [0.673;0.717] | 0.363 [0.327;0.396] | 0.915 [0.895;0.939] | 0.00 |
| DCM | 83 | 64 | 0.843 [0.822;0.858] | 0.872 [0.844;0.892] | 0.630 [0.600;0.650] | 0.888 [0.875;0.892] | 0.595 [0.545;0.632] | 0.00 |
| Myocarditis | 46 | 9 | 0.902 [0.842;0.958] | 0.884 [0.778;1.000] | 0.849 [0.789;0.895] | 0.586 [0.500;0.692] | 0.969 [0.938;1.000] | 0.00 |
| HCM | 60 | 6 | 0.945 [0.861;0.988] | 0.978 [0.833;1.000] | 0.599 [0.574;0.630] | 0.214 [0.179;0.231] | 0.996 [0.969;1.000] | 0.00 |
| Ischemia | 81 | 8 | 0.902 [0.864;0.938] | 0.894 [0.857;1.000] | 0.827 [0.797;0.851] | 0.354 [0.300;0.400] | 0.987 [0.983;1.000] | 0.00 |
| Valvular Disease | 42 | 2 | 0.872 [0.839;0.901] | 0.500 [0.500;0.500] | 0.839 [0.800;0.854] | 0.135 [0.111;0.143] | 0.971 [0.970;0.972] | 0.00 |
| Congenital Heart Disease | 43 | 1 | 0.832 [0.810;0.857] | 1.000 [1.000;1.000] | 0.733 [0.714;0.762] | 0.082 [0.077;0.091] | 1.000 [1.000;1.000] | 0.00 |
| **Utrecht** | 1171 | 206 | 0.833 [0.820;0.844] | 0.894 [0.874;0.913] | 0.555 [0.542;0.567] | 0.300 [0.292;0.308] | 0.961 [0.953;0.968] | 0.00 |
| Male | 747 | 157 | 0.843 [0.828;0.857] | 0.910 [0.886;0.930] | 0.549 [0.533;0.566] | 0.349 [0.338;0.360] | 0.958 [0.948;0.968] | 0.00 |
| Age <60 | 529 | 65 | 0.811 [0.789;0.830] | 0.857 [0.815;0.892] | 0.557 [0.538;0.574] | 0.214 [0.203;0.225] | 0.965 [0.955;0.974] | 0.00 |
| Age ≥60 | 638 | 140 | 0.842 [0.827;0.857] | 0.911 [0.885;0.936] | 0.553 [0.536;0.570] | 0.363 [0.352;0.375] | 0.957 [0.945;0.968] | 0.00 |
| QRS <100 ms | 625 | 52 | 0.832 [0.799;0.861] | 0.837 [0.776;0.891] | 0.646 [0.630;0.664] | 0.177 [0.158;0.196] | 0.978 [0.969;0.984] | 0.00 |
| QRS ≥100 ms | 545 | 154 | 0.786 [0.768;0.803] | 0.914 [0.890;0.935] | 0.421 [0.401;0.441] | 0.382 [0.367;0.395] | 0.926 [0.905;0.945] | 0.00 |
| Heart Rate >80 bpm | 301 | 75 | 0.822 [0.795;0.847] | 0.919 [0.882;0.957] | 0.454 [0.425;0.484] | 0.358 [0.327;0.389] | 0.944 [0.918;0.970] | 0.00 |
| Heart Rate ≤80 bpm | 869 | 131 | 0.831 [0.813;0.848] | 0.880 [0.848;0.909] | 0.585 [0.571;0.601] | 0.273 [0.258;0.287] | 0.965 [0.956;0.973] | 0.00 |
| Atrial Fibrillation | 50 | 21 | 0.852 [0.775;0.919] | 0.989 [0.947;1.000] | 0.310 [0.210;0.414] | 0.507 [0.436;0.579] | 0.975 [0.875;1.000] | 0.00 |
| No Atrial Fibrillation | 1120 | 185 | 0.828 [0.815;0.841] | 0.883 [0.859;0.906] | 0.562 [0.550;0.575] | 0.285 [0.276;0.294] | 0.961 [0.953;0.968] | 0.00 |
| Coronary Disease | 320 | 82 | 0.825 [0.802;0.846] | 0.919 [0.889;0.951] | 0.527 [0.498;0.559] | 0.401 [0.381;0.420] | 0.950 [0.932;0.968] | 0.00 |
| Other CMP | 139 | 27 | 0.797 [0.762;0.828] | 0.913 [0.849;0.963] | 0.493 [0.469;0.522] | 0.299 [0.275;0.321] | 0.960 [0.932;0.983] | 0.00 |
| DCM | 83 | 64 | 0.745 [0.711;0.772] | 0.866 [0.828;0.906] | 0.366 [0.350;0.368] | 0.821 [0.809;0.829] | 0.451 [0.389;0.538] | 0.00 |
| Myocarditis | 46 | 9 | 0.845 [0.778;0.919] | 0.950 [0.889;1.000] | 0.446 [0.368;0.526] | 0.292 [0.258;0.333] | 0.974 [0.933;1.000] | 0.00 |
| HCM | 60 | 6 | 0.742 [0.656;0.781] | 0.994 [0.833;1.000] | 0.377 [0.333;0.417] | 0.151 [0.132;0.158] | 0.998 [0.955;1.000] | 0.00 |
| Ischemia | 81 | 8 | 0.922 [0.847;0.966] | 0.952 [0.750;1.000] | 0.565 [0.514;0.603] | 0.188 [0.150;0.216] | 0.991 [0.955;1.000] | 0.00 |
| Valvular Disease | 42 | 2 | 0.774 [0.750;0.805] | 0.500 [0.500;0.500] | 0.681 [0.625;0.732] | 0.072 [0.062;0.083] | 0.965 [0.962;0.968] | 0.00 |
| Congenital Heart Disease | 43 | 1 | 0.853 [0.810;0.881] | 1.000 [1.000;1.000] | 0.524 [0.524;0.524] | 0.048 [0.048;0.048] | 1.000 [1.000;1.000] | 0.00 |

Abbreviations: AUROC: area under the receiver operating curve, CI: confidence interval, PPV: positive predictive value, NPV: negative predictive value, CGMH: Chang Gung Memorial Hospital.

**Supplementary table 11.** Recalibration of model thresholds

| Model | Threshold | N patients | N outcome | Mean Threshold | AUC | Sensitivity | Specificity | PPV | NPV | Number missing |
| --- | --- | --- | --- | --- | --- | --- | --- | --- | --- | --- |
| AiTiALVSD | Original | 1203 | 211 | 0.119 | 0.938 [0.934; 0.942] | 0.882 [0.864; 0.904] | 0.855 [0.847; 0.863] | 0.565 [0.552; 0.579] | 0.971 [0.967; 0.976] | 0.00 |
| AiTiALVSD | Youden | 1203 | 211 | 0.096 | 0.938 [0.934; 0.942] | 0.908 [0.878; 0.933] | 0.839 [0.810; 0.872] | 0.548 [0.510; 0.594] | 0.977 [0.971; 0.983] | 0.00 |
| CGMH | Original | 1203 | 211 | 0.450 | 0.902 [0.893; 0.910] | 0.843 [0.817; 0.872] | 0.809 [0.800; 0.819] | 0.486 [0.469; 0.500] | 0.960 [0.954; 0.967] | 0.00 |
| CGMH | Youden | 1203 | 211 | 0.417 | 0.902 [0.893; 0.910] | 0.866 [0.817; 0.915] | 0.802 [0.747; 0.839] | 0.485 [0.435; 0.526] | 0.966 [0.955; 0.977] | 0.00 |
| Utrecht | Original | 1129 | 192 | 0.105 | 0.832 [0.821; 0.844] | 0.890 [0.870; 0.910] | 0.558 [0.546; 0.569] | 0.293 [0.283; 0.304] | 0.961 [0.955; 0.968] | 6.09 |
| Utrecht | Youden | 1129 | 192 | 0.307 | 0.832 [0.821; 0.844] | 0.757 [0.712; 0.822] | 0.778 [0.714; 0.819] | 0.415 [0.366; 0.453] | 0.940 [0.931; 0.952] | 6.09 |
| ECG Vision | Original | 1203 | 211 | 0.100 | 0.886 [0.877; 0.896] | 0.814 [0.796; 0.830] | 0.817 [0.808; 0.824] | 0.487 [0.473; 0.497] | 0.954 [0.949; 0.958] | 0.00 |
| ECG Vision | Youden | 1203 | 211 | 0.100 | 0.886 [0.877; 0.896] | 0.817 [0.760; 0.872] | 0.816 [0.765; 0.857] | 0.489 [0.445; 0.543] | 0.955 [0.943; 0.966] | 0.00 |

Abbreviations: AUROC: area under the receiver operating curve, CI: confidence interval, PPV: positive predictive value, NPV: negative predictive value, CGMH: Chang Gung Memorial Hospital.

**Supplementary table 12.** False negative and positive percentage rate per LVEF category

| Model | LVEF Category | Total ECGs | False Positive % | False Positives | False Negative % | False Negatives |
| --- | --- | --- | --- | --- | --- | --- |
| CGMH | LVEF 40-50% | 938 | 52.77186 | 495 |  |  |
| CGMH | LVEF >50% | 2522 | 15.26566 | 385 |  |  |
| CGMH | LVEF <40% | 1277 |  |  | 14.40877 | 184 |
| ECG Vision | LVEF 40-50% | 938 | 56.28998 | 528 |  |  |
| ECG Vision | LVEF >50% | 2522 | 17.16891 | 433 |  |  |
| ECG Vision | LVEF <40% | 1277 |  |  | 14.33046 | 183 |
| AiTiALVSD | LVEF 40-50% | 938 | 52.87846 | 496 |  |  |
| AiTiALVSD | LVEF >50% | 2522 | 11.69707 | 295 |  |  |
| AiTiALVSD | LVEF <40% | 1277 |  |  | 10.25842 | 131 |
| Utrecht | LVEF 40-50% | 938 | 64.60554 | 606 |  |  |
| Utrecht | LVEF >50% | 2522 | 39.96828 | 1008 |  |  |
| Utrecht | LVEF <40% | 1277 |  |  | 8.222396 | 105 |

Abbreviations: LVEF: left ventricular ejection fraction
